# Molecular evolution of Andes virus lineages leading to recent human outbreaks

**DOI:** 10.64898/2026.11.06.26367422

**Authors:** Charu Sharma, Sanni Översti, Robert Stass, Alexander G Lucaci, Jonathan Audet, David Safronetz, Lele Zhao, Katrina Lythgoe, Aris Katzourakis, Thomas A. Bowden, Spyros Lytras, Mahan Ghafari

## Abstract

Andes virus (ANDV) is exceptional among hantaviruses in its capacity for person-to-person transmission, but whether outbreaks reflect recent viral adaptation or emergence of pre-existing reservoir lineages remains unclear. We generated paired blood- and nasal-derived genomes from the MV Hondius outbreak and integrated them with a segment-linked collection of human- and rodent-derived ANDV sequences, including incomplete genomes. We developed a linkage-informed missing-segment framework that preserves isolate identity, sampling time, and cross-segment placement without imputing nucleotide states, enabling sensitivity analyses of patchy segment coverage. Across five human outbreaks, dated phylogenies placed outbreak lineages independently within geographically structured reservoir diversity, with closest sampled ancestry predating detection. Time-dependent-rate analyses indicated a deeper reservoir-associated history, while mutation reconstruction, selection tests, branch-rate comparisons, and structural mapping found no recurrent adaptive transition preceding human transmission. MV Hondius samples showed minimal consensus-level diversification. These results shift attention from repeated recent adaptation toward deeper lineage properties and ecological opportunity.

## Introduction

Andes hantavirus (ANDV) causes severe hantavirus cardiopulmonary syndrome and is unusual among hantaviruses in its capacity for documented person-to-person transmission^1^. ANDV has a tripartite negative-sense single-stranded RNA genome in which the S segment encodes the nucleoprotein, the M segment encodes the glycoprotein precursor that gives rise to Gn and Gc, and the L segment encodes the RNA-dependent RNA polymerase (RdRp)^2^. Historical sequencing is uneven across these segments, with many isolates represented by only one or two segments and some M-segment records available only as partial Gn/Gc sequences with separate accessions. ANDV circulates primarily in the long-tailed colilargo (*Oligoryzomys longicaudatus*), with occasional detection in other rodents (e.g. *Loxodontomys micropus, Abrothrix longipilis*), and repeatedly spill over into humans in Argentina and Chile^3,4^. The 2018-2019 Epuyén outbreak^5^ and the 2026 MV Hondius-associated outbreak have therefore raised a central question: are ANDV outbreaks involving person-to-person transmission driven by the recent adaptive viral changes, or by the ecological emergence of pre-existing reservoir lineages already capable of limited human transmission? Previous comparative work proposed candidate mutations associated with person-to-person transmission^6^, but whether independent outbreaks share a recurrent recent adaptive transition has not been tested systematically.

These alternatives make different evolutionary predictions. Under a model of recent adaptive emergence, independently arising human outbreaks should repeatedly be preceded by one or more signatures of viral adaptation. These may include features such as accelerated nonsynonymous evolution, signals of positive selection on outbreak-lineage ancestral branches, convergent amino acid changes, or recurrent perturbation of viral structures involved in entry, replication, immune evasion, or antigenicity. Under an ecological-emergence model, outbreak lineages should instead be distributed across standing reservoir diversity, retain strong geographic structure, and lack a reproducible genomic transition immediately before their detection in humans.

For many emerging zoonoses, genomic epidemiology focuses on reconstructing transmission chains after detection in humans. For reservoir-borne viruses, however, the deeper question is often whether the human outbreak lineage can be placed within known animal diversity and whether the branch leading to human emergence shows evidence of accelerated or adaptive evolution.

This is particularly challenging for ANDV, for which available genomic data are sparse and unevenly sampled across human and rodent hosts^7^. Available sequences include known human outbreak clusters, rodent-associated genomes, and ANDV-like sequences that have not always been consistently classified within major clades. Historical isolates are also frequently represented by incomplete segment sets, resulting in substantial differences in sequence availability across segments and sampling dates. These characteristics can complicate molecular dating, as uneven temporal and host sampling can distort the available temporal signal, while incomplete segment sampling and uncertainty in clade classification can influence the inferred phylogenetic structure.

Together, these factors may affect evolutionary-rate and divergence-time estimates, and potentially bias inferences about outbreak origins.

Here, we use the MV Hondius 2026 outbreak as an entry point into a broader comparative analysis of ANDV evolution. We generated a paired blood- and nasal-derived S, M, and L segment sequence from the outbreak and integrated them with a segment-linked collection of public human, rodent, and ANDV-like sequences, including incomplete and partial genomes. We first reconstruct population-level diversity and introduce a linkage-informed missing-segment framework for retaining sampled viruses in segment alignments when a linked segment is unavailable. We then systematically infer molecular rate estimates, evaluate the presence of time-dependent rate variation across nested datasets and apply the Prisoner of War (PoW) model^8^ to examine deeper evolutionary timelines of ANDV. Against this macroevolutionary background, we compare the evolutionary emergence of MV Hondius outbreak with four earlier outbreaks involving documented human-to-human transmission: i) a household and nosocomial transmission chain in Southern Chile with five human infections in 2011 (Chile 2011)^9^, ii) a three-person transmission cluster in Argentina in 2014 (Argentina 2014)^10^, iii) the largest known ANDV outbreak with 34 confirmed infections in Argentina in 2018-2019 (Epuyén 2018/19)^5^, and iv) a two-person family cluster from Chile in 2024 (Chile 2024)^11^. We then identify the closest sampled relatives and geographic context of each outbreak lineage, reconstruct their evolutionary ancestry, and identify coding substitutions that arose along the ancestral branch leading to each outbreak. By combining these reconstructions with branch-rate, codon-selection, and structural analyses, we directly test whether repeated human outbreaks share evidence of recent adaptive emergence while explicitly accounting for the patchiness of the available genomic records.

## Results

### Phylogenetic placement and molecular dating of recorded ANDV human outbreaks

To best characterise the broader diversity that ANDV sits in, we first reconstructed maximum-likelihood phylogenies for all publicly available S, M and L genome segments of the *Orthohantavirus andesense* (**Supplementary Figures 1-3**). Consistent with previous studies^6^, the five recorded human-to-human outbreak lineages fell within the four main ANDV clades and were embedded within the broader rodent-associated ANDV diversity. Our expanded curation additionally retains incomplete and partial genome records, including rodent-derived isolates and the Chile 2011 human outbreak ^4,9^ that would be lost by restricting analysis to complete three-segment genomes, thereby broadening the reservoir context available for each outbreak. The closest sampled relatives of the outbreak lineages were regionally structured. MV Hondius 2026 and Chile 2024 clustered within ANDV clade 3, together with rodent-derived (*O. longicaudatus*) sequences^4^ and isolated human-infection sequences from Valdivia, Los Ríos, and San Martín de los Andes, Neuquén, together with related Neuquén sequences^9^. This pattern is consistent with a cross-border southern Chile-northern Patagonia assemblage rather than a source confined to one national surveillance system (**Figure 1A**, **Supplementary Figures 1-3)**. The Argentina 2014 and Epuyén 2018/19 lineages were placed within ANDV clade 2, whereas Chile 2011 fell within clade 1. No human-to-human transmission has been documented yet for members of ANDV clade 4 (which contains the reference ANDV genome, strain Chile-9717869^2^), although it contains genomes derived from isolated human infections. Several rodent-derived sequences fell within ANDV diversity but outside clades 1-4. In particular, two well-supported groups were recovered in M and S segments. Several other isolates also occupied positions outside clades 1-4, but these placements were more weakly supported and/or inconsistent among genome segments. These sequences may represent additional, previously under-recognised lineages of ANDV. We refer to all of these sequences and clusters as “unclassified” (**Supplementary Figures 1-3)**.

**Figure 1.**
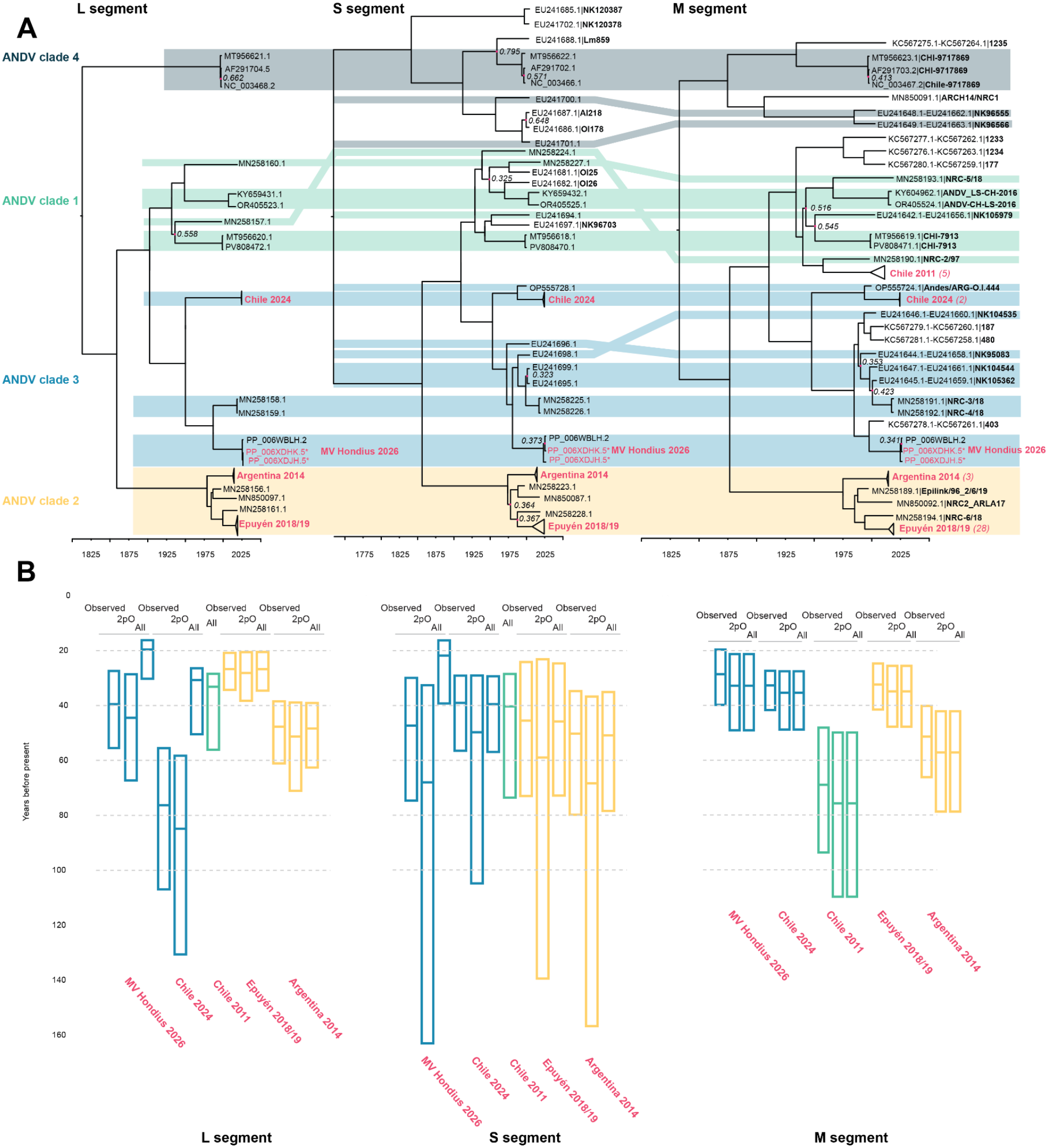
Time-scaled ancestry of the five documented ANDV human outbreaks across genome segments. **(A)** Dated maximum clade credibility trees for the L, S, and M coding sequences. Genomes with sequences available for multiple segments are connected between the phylogenies and coloured based on the ANDV clade they are part of. Sequences of the MV Hondius 2026 outbreak reported in this study are highlighted in pink and followed by an asterisk. Nodes with posterior support below 0.8 are annotated with pink dots and their posterior support value is shown next to them. **(B)** Posterior distributions for the closest sampled common ancestor of each outbreak lineage under three sampling configurations: observed-sequence-only datasets (“Observed”), observed-sequence datasets retaining two representatives per outbreak (“2pO”), and linkage-informed datasets containing missing-segment taxa (“Linkage-informed”; labelled “All” in the panel). Horizontal lines show posterior medians and box limits show 95% highest posterior density intervals.

The nearest sampled relatives of each outbreak lineage differed among genome segments, owing to uneven sequence availability across the segmented genome (**Figure 1**). We refer to these non-outbreak human- or rodent-derived nearest relatives as comparator sequences, because they provide the phylogenetic context for placing outbreak lineages within broader ANDV diversity. The M segment contained the richest set of nearby comparators for all outbreak lineages (total of 78 available ANDV M segment sequences), followed by the S segment (70 available ANDV sequences). The L segment had fewer nearby comparators and lacked equivalent closely related rodent-derived sequences, limiting reservoir-inclusive inference (50 available ANDV sequences). These differences in segment availability are central to the molecular dating problem addressed below: although sequences from different segments may be linked to the same isolate, observed-sequence-only alignments cannot represent that linkage when one or more segments are missing.

Analysing only taxa with observed sequence data in each segment therefore produced segment-specific datasets with different taxonomic compositions and, for a small subset of clades, poorly resolved or unstable time trees. Rather than discarding incompletely sequenced isolates, we developed a linkage-informed missing-segment representation to retain known linkage without imputing sequence states (see **Methods**). If a sampled virus had an observed sequence for at least one segment but lacked another, the missing segment was represented by a genetically non-informative sequence (i.e., an all-N sequence of the appropriate length) and assigned the same isolate identity and sampling date. Its placement was constrained to the clade or internal relationship supported by the linked observed segment(s), while unconstrained relationships remained estimated from the segment-specific data. The approach therefore carries known linkage, sampling time, and explicit cross-segment topology assumptions; it does not reconstruct the missing sequence, and its assumptions can be violated by reassortment or uncertainty in the constrained topology. We accordingly interpret linkage-informed estimates alongside the observed-sequence dataset.

Observed-sequence S, M, and L trees placed outbreak-associated lineages and their nearest comparators in broadly congruent positions (**Figure 1A**). Because missing-segment taxa are constrained by linked segments, we assessed reassortment only in observed-sequence trees. We found no well-supported segment discordance involving the five outbreak lineages or their closest comparators, with the caveat that only M-segment data are available for Chile 2011. Few placements differed among segments, but most involved weakly supported nodes (posterior probability <0.8). Thus, reassortment may have partly contributed to ANDV evolution, but it does not affect the placement of the recent outbreak lineages analysed here.

### Molecular clock behaviour and deeper evolutionary history of ANDV

We next systematically evaluated temporal signal and time-dependent rate variation across nested ANDV datasets. For each genome segment, we analysed progressively broader phylogenetic subsets comprising clade 2 alone; a combined clade 1 + clade 3 dataset containing only clades 1 and 3; clades 1-3 containing clades 1, 2 and 3; and clades 1-4 containing all four major ANDV clades. Bayesian Evaluation of Temporal Signal (BETS)^12^ compared heterochronous models incorporating the observed sampling dates with otherwise equivalent isochronous models in which all tips were assigned the same date (**Methods**). BETS supported the presence of temporal signal for all M-segment datasets, and most S-segment datasets (except the clade1 + clade 3 datasets). Temporal signal was weaker for the L-segment, consistent with fewer sequences, lacking signal for clade 2, clade 1 + clade 3, and clade 1-3 datasets (**Supplementary Table 4**). However, the inclusion of linkage-informed missing-segment taxa and the associated constraints on the tree topology stabilised the time-scaled phylogeny, produced a consistent rate estimate across independent runs, and restored support for temporal signal by BETS (see **Supplementary Table 4**). Because the missing-segment taxa add no nucleotide states, this gain should be interpreted as improved use of known sampling and linkage information rather than additional sequence evidence. This improvement was particularly important for the more sparsely sampled L segment, where incomplete taxon representation otherwise limited time-tree inference.

Across the nested datasets, posterior evolutionary rate estimates declined as older and more divergent ANDV diversity was incorporated (**Figure 2A**, **Supplementary Table 5**). Clade 2 tended to produce the highest rate estimates, whereas broader clade combinations produced lower estimates. This systematic decline in inferred rate with increasing phylogenetic depth is consistent with time-dependent rate variation. Across different sampling configurations, rate estimates were comparatively stable except where topology uncertainty was substantial, as in L clades 1+3. By contrast, closest-sampled-ancestor dates were more sensitive to comparator representation (**Figure 1B**). The principal value of the linkage-informed analysis is therefore not added sequence evidence or guaranteed precision, but explicit representation and testing of known cross-segment linkage and topology assumptions under patchy segment coverage.

**Figure 2.**
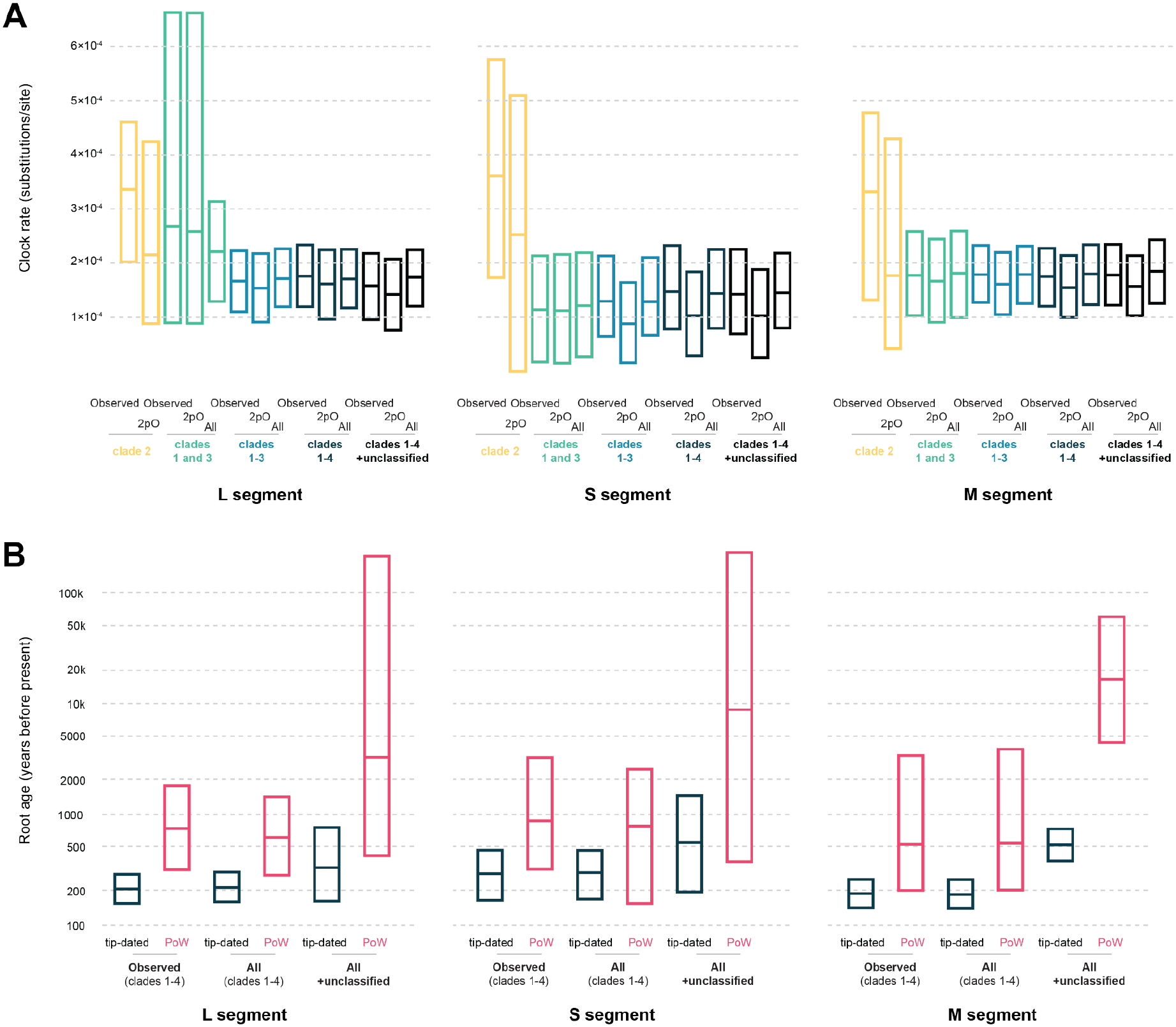
Time-dependent rates and deep evolutionary reconstruction. **(A)** Posterior strict-clock rate distributions for nested ANDV datasets across L, S, and M segments. “Observed” denotes observed-sequence-only datasets; “2pO” denotes observed-sequence datasets with two outbreak samples retained; and “All” denotes linkage-informed datasets. **(B)** Posterior root-age distributions for tip-dated and PoW-transformed trees under three configurations: observed clades 1-4, linkage-informed clades 1-4, and linkage-informed clades 1-4 plus unclassified lineages. Root ages are shown on a log10 years-before-present scale. Horizontal lines indicate medians and box limits indicate 95% HPD intervals.

Time-dependent rate variation is well described in RNA viruses, with short-term evolutionary-rate estimates often exceeding those inferred over deeper timescales^13^. In RNA viruses, this can arise because recent datasets retain transient polymorphisms and mildly deleterious mutations that are later removed by purifying selection^8^. This interpretation is also consistent with our codon-based analyses, which indicate that purifying selection dominates most coding regions of the ANDV genome. In ANDV, the effect may be particularly apparent because older pre-outbreak human- and rodent-derived sequences can remain genetically very similar to much more recent outbreak lineages. Incorporating these older comparator sequences therefore increases the temporal depth of the dataset without a proportional increase in genetic divergence, which lowers the substitution rate required to explain the observed sequence differences. Thus, as progressively older reservoir-associated diversity is included, both the sampled phylogenetic depth and the timescale over which the molecular clock is estimated increase, producing the lower rate estimates observed in the broader clade datasets.

We therefore treated the tip-dated analyses as estimates of sampled ancestry over recent-to-intermediate timescales, rather than as direct estimates of the deeper age of ANDV clades. To explore deeper timescales, we applied the Prisoner of War (PoW) model, which accounts for the decline in apparent substitution rates over longer evolutionary intervals (**Methods**). We used the clade 2 inferred rate as the calibration for the PoW model over short timescales.

Across all three genome segments, PoW-transformed root-age estimates were older than the corresponding tip-dated estimates (**Figure 2B**). For the observed clades 1-4 datasets and the corresponding linkage-informed clades 1-4 datasets, however, the two approaches showed broadly similar patterns: PoW shifted median root ages deeper in time, but the 95% HPD intervals generally overlapped those of the tip-dated analyses. The larger change occurred when the additional unclassified ANDV diversity was included. Across segments, the clades 1-4 + unclassified datasets produced older root-age estimates, although the magnitude and precision of this shift depended strongly on the genomic information available for these divergent lineages.

For the S and L segments, several of the unclassified isolates were represented by linkage-informed missing-segment taxa rather than observed nucleotide sequences. Because these all-N taxa contribute no nucleotide-state information, their inclusion allows known isolate linkage, sampling time and phylogenetic placement to be represented without imputing sequence evolution but consequently leaves substantial uncertainty in the amount of divergence associated with these deeper branches. This is reflected in the very broad 95% HPD intervals of the S- and L-segment clades 1-4 + unclassified PoW estimates. In contrast, the M segment contains the largest set of observed and relatively divergent unclassified sequences and therefore provides substantially more direct sequence information about this deeper diversity. Incorporating these sequences shifted the inferred M-segment root markedly deeper, to approximately 16,500 years before present (95% HPD: 4,500-60,500 years). Although this interval remains broad and should not be interpreted as a precise estimate of the origin of ANDV, the comparatively well-resolved M-segment analysis supports the possibility that the sampled ANDV lineage extends back many thousands of years, substantially predating the recent human outbreaks through which much of its diversity was first recognised.

### Linkage-informed dating of individual outbreak lineages

We compared the closest sampled common-ancestor dates of each outbreak lineage across three dataset configurations: the observed-sequence datasets, linkage-informed datasets containing missing-segment taxa, and reduced observed-sequence datasets containing two representative sequences per outbreak cluster (“2pO”) (**Figure 1B**, **Supplementary Table 6**). For MV Hondius 2026, the M segment contained the nearest sampled comparator, strain 403 from Valdivia, Chile (2010; Gn KC567278.1, Gc KC567261.1). In the observed-sequence analysis, the median closest sampled common-ancestor date was 1997 (95% HPD: 1986-2006). The S and L segments gave older median estimates of 1979 (95% HPD: 1951-1996) and 1986 (95% HPD: 1970-1998), respectively, but lacked equivalent closely related comparators. Including linkage-informed missing-segment taxa allowed the same known comparator lineages to be represented more consistently across segments and produced a more coherent set of segment-specific estimates, with inferred dates of 2004 (95% HPD: 1987 - 2010), 1998 (95% HPD: 1986 - 2006), and 2006 (95% HPD: 1996 - 2010) for the S, M, and L segments, respectively. The missing-segment-taxa analysis should therefore be read as a sensitivity analysis of how known but incompletely sequenced taxa alter molecular dating.

For Epuyén 2018/19 lineage, the closest sampled common ancestor was dated to 1980 (95% HPD: 1953 - 2001) for S, 1994 (95% HPD: 1985 - 2002) for M and 1999 (95% HPD: 1991 - 2005) for L; the closest sampled outgroup were also from Argentina^5^. Because segment coverage for the relevant clade 2 comparators was already relatively complete, linkage-informed analyses produced little change in these estimates. By contrast, the Chile 2024 lineage highlights the effect of uneven segment coverage. For S and M, where the closest comparator was already represented, observed and linkage-informed estimates were nearly identical (S: 1987 vs. 1986; M: 1993 vs. 1993). For L, the corresponding comparator lacked sequence data, and the estimate shifted from 1950 to 1995 when represented as a linkage-informed missing-segment taxon, bringing it into much closer agreement with S and M (**Figure 1B**). This shows how cross-segment linkage can materially improve dating when a close comparator is missing from one segment.

The remaining reported human outbreak lineages were similarly placed and dated within their nearest available segment-specific comparator clades, with estimates varying according to the density and segment coverage of available comparator sequences (**Figure 1B**). Argentina 2014 lineage fell as an outgroup to Epuyén 2018/19 lineage within the Río Negro- and Chubut-associated diversity; Chile 2011 lineage clustered near Río Negro-associated diversity, and Chile 2024 lineage clustered near Neuquén-associated diversity. The 2pO analyses indicated that dates were not driven by dense sampling of near-identical outbreak genomes (**Figure 1B**). These dates describe the closest sampled ancestral relationship, not spillover timing, the first human infection, or geographic origin. Because reservoir sampling remains sparse, decade-scale gaps to comparators are best interpreted as unsampled reservoir ancestry.

### No shared recent adaptive transition precedes human outbreaks

Because the five documented outbreaks form distinct lineages within the ANDV phylogeny, we asked whether independent emergences were repeatedly preceded by common evolutionary signatures. We reconstructed coding changes along the branch connecting the closest ancestral node shared with sampled comparators to the common ancestor of each outbreak lineage (the outbreak stem). For each segment, ancestral states were reconstructed in local clades containing outbreak sequences and their nearest sampled relatives.

Across the five outbreak stems, most changes were synonymous, with only a few nonsynonymous changes in M and L (MV Hondius 2026: L, 49 synonymous; S, 12 synonymous; M, 17 synonymous and 1 nonsynonymous. Epuyén 2018/19: L, 19 synonymous; S, 6 synonymous; M, 15 synonymous. Chile 2024: L, 85 synonymous and 2 nonsynonymous; S, 4 synonymous; M, 23 synonymous. Argentina 2014: L, 31 synonymous and 2 nonsynonymous; S, 6 synonymous; M, 16 synonymous. Chile 2011: M, 6 synonymous and 3 nonsynonymous). Only eight nonsynonymous changes were reconstructed across all available outbreak-lineage ancestral branches, and no amino acid substitution recurred consistently across the five emergences (**Supplementary Table 7**). Neither the number nor distribution of nonsynonymous substitutions supported a general burst of protein-changing evolution immediately before human outbreak detection.

To assess whether any nonsynonymous substitution had an obvious mechanistic interpretation, we combined site-specific selection analyses with structural mapping to the available ANDV glycoprotein structure (PDB 9P3I^14^) and homologous Hantaan virus RdRp structures (PDB 8P1L and 5IZE^15,16^) (**Figure 3**). We used structural mapping to generate mechanistic hypotheses, not to infer phenotypic effects in the absence of experimental evidence.

**Figure 3.**
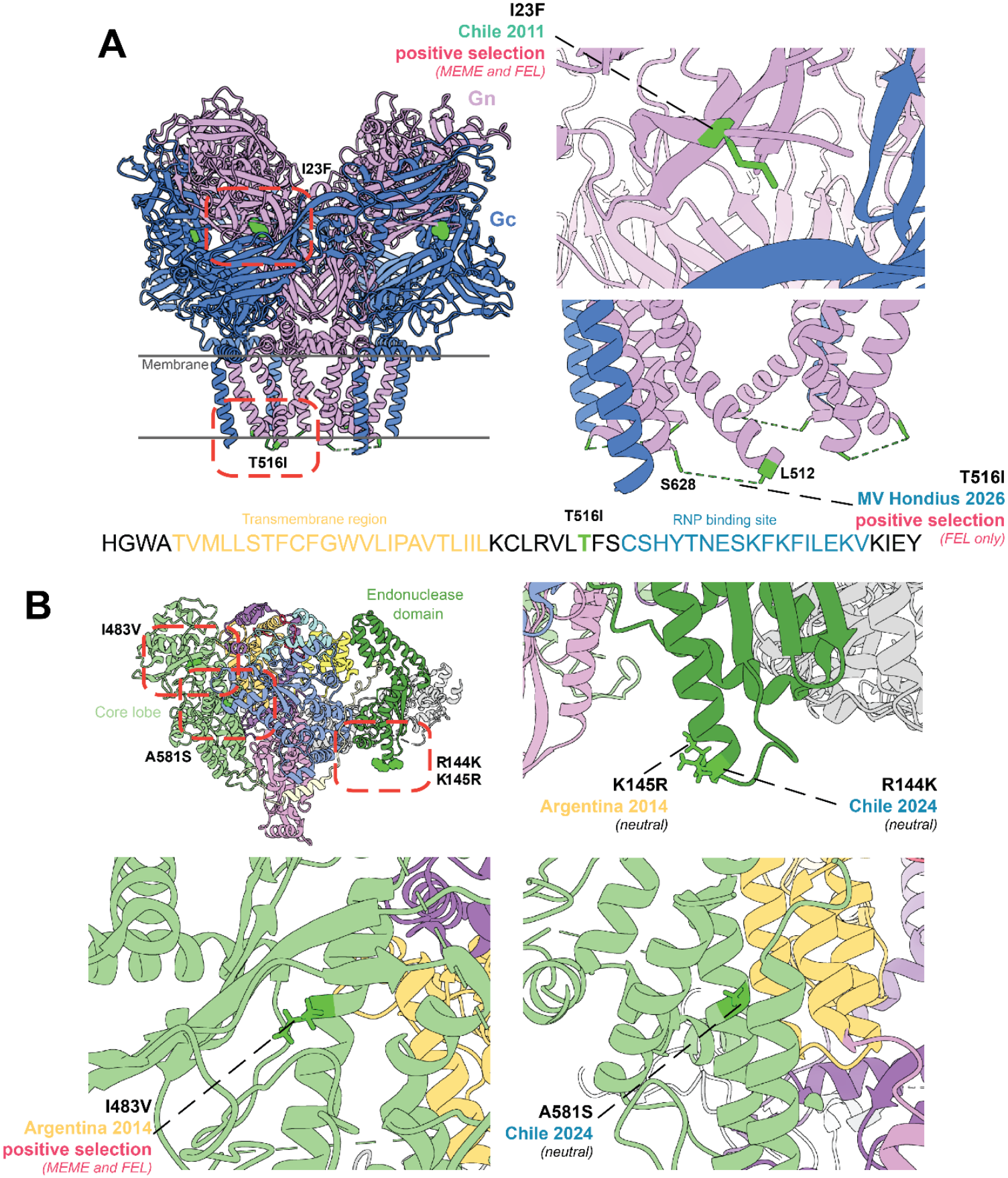
Structural context of nonsynonymous substitutions detected in the stem branch of outbreak-associated ANDV lineages. **(A)** ANDV glycoprotein complex cryoEM structure (PDB ID 9P3I). A segment of sequence is displayed to show the proximity of T516I to a transmembrane region at residues 486 to 509 and a putative RNP (N protein) binding site between residues 519 and 536. **(B)** Hantaan virus polymerase complex cryoEM structure (PDB ID 8P1L), used in the absence of an Andes polymerase structure. These corresponding sites are L144, R145, I482 and A580 respectively on the Hantaan polymerase. The colour scheme for the RdRp structure matches that used in the associated manuscript^15^. Both structures are colored by their subunits and sites of interest are highlighted in green. Zoomed in views of each region of interest (highlighted with red dashed line rectangles on the full structure) are shown on the right (A and B) and bottom left (B) of each panel. The substitution, outbreak lineage on the stem branch of which it took place, and selection results for the two site-specific selection methods FEL and MEME are annotated on each view.

The M segment encodes the glycoprotein precursor, which is cleaved into Gn and Gc, mediates entry, and presents major neutralising-antibody targets^17^. The MV Hondius stem carried T516I in a linker between the transmembrane region and a proposed RNP/N-protein-binding region^18,19^ (**Figure 3A**). The substitution changes a polar residue to a hydrophobic residue adjacent to L515 and F517. FEL provided only weak, non-significant evidence of diversifying selection (p=0.09), and the site does not map to an exposed antigenic surface in the prefusion structure; any effect on RNP binding remains unresolved. Chile 2011 carried I23F at the N terminus of the Gn head domain, directed towards a groove on Gc. Although residue 23 is solvent exposed in the isolated domain, it is inaccessible to antibodies in the prefusion assembly. FEL and MEME likewise provided only weak, non-significant evidence at this site (p=0.06 and p=0.09, respectively). The additional Chile 2011 substitutions V905I and V997I showed no selection signal or obvious structural mechanism.

A similar number of stem branch amino acid changes were detected on the polymerase (L segment). Two of these are structurally adjacent to one another but took place on the stem branch of different human outbreak lineages: R144K in Chile 2024, and K145R in Argentina 2014 (**Figure 3B**). These mutations are found at the tip of the αd helix in the RdRp endonuclease domain. This helix is extended in hantaviruses relative to other viral families, so the function of this region is unknown and does not form part of the endonuclease active site^20^. The mutations are like-for-like basic substitutions that are unlikely to affect protein functionality and exhibit no detectable signal of selection. However, given that these mutations occurred independently in the stem branch of two ANDV human outbreaks, we cannot discount the possibility of phenotypic relevance. Polymerase amino acid changes were I483V (Argentina 2014) and A581S (Chile 2024) were also observed in the RdRp core lobe. I483V is located on the viral RNA binding lobe (vRBL)^16^ and A581S to the interface with the RdRp thumb domain (**Figure 3B**). Both sites are buried within the protein core and do not directly interact with vRNA. Although these mutations do not introduce an obvious functional effect, I483V is detected to be under positive selection by both methods used in the analysis (FEL p-value = 0.04; MEME p-value = 0.06).

Across the broader phylogeny, negative selection dominated most coding regions. Branch-level tests showed no evidence for episodic diversifying selection on any of the five outbreak stems. Relaxed clock branch rate analyses likewise did not identify elevated rates on outbreak stems (**Supplementary Figure 4**). L-segment relaxed clock estimates were broader than strict clock estimates, but outbreak-stem estimates remained within background distributions. We therefore found no evidence that outbreak emergence was preceded by a general acceleration in molecular evolutionary rate.

Lastly, we examined selection signatures on the stems of ANDV clades 1-4. RELAX provided little evidence for widespread shifts in selection intensity across L, M, and S. Only the S-segment comparison showed significant intensification (K=2.05, p=0.015); other comparisons were neutral or showed non-significant trends. Any deeper shift in selective pressure therefore appears segment-specific rather than genome-wide, with the strongest evidence involving S.

### Within-host diversity in the MV Hondius 2026 outbreak

We generated two primary S, M, and L segment sequence sets from the MV Hondius 2026 outbreak, using paired whole-blood and nasopharyngeal samples collected from the same individual on the same day (see **Methods**), and analysed them alongside the previously reported outbreak genome (PP_006WBLH.2). Across these three consensus genomes, diversity was extremely limited; the two newly generated consensus sequences differed by one synonymous substitution in the L segment, L:C2144T. The previously reported outbreak genome contained one additional synonymous L-segment difference^21^, further indicating that consensus-level diversity remained extremely limited during the sampled outbreak.

We next examined whether low-frequency within-host variation captured additional evolutionary diversity not apparent from consensus genomes. After filtering for coverage and depth (see **Methods**), several iSNVs were independently detected in both blood and nasal samples from the same individual. These included three synonymous variants, S:T909G, M:A1164G and L:G407A, and two nonsynonymous variants, S:G916A, which changes alanine to threonine at amino acid position 292 of the S segment, and L:T3561C, which changes phenylalanine to leucine at amino acid position 1176 of the L segment. Across the paired samples, variant frequencies ranged from 5.1% to 23.4%, with at least 20× read depth at each position (**Supplementary Figure 5** and **Supplementary Table 8**).

Most shared iSNVs remained at low frequency and did not correspond to fixed differences among the MV Hondius outbreak genomes, consistent with limited diversification during the observed transmission chain. However, several within-host variable positions also overlapped sites reconstructed as substitutions along the outbreak-lineage ancestral branches of one or more of the five human outbreaks, including S: C1116T; L:A704G, L:T3560C, L: T6302C, and M:G1164A (see **Supplementary Tables 7 and 8**). Together, these findings show that the MV Hondius outbreak contained greater low-frequency diversity than was visible from consensus genomes, while providing no evidence that a new variant rose to fixation during the sampled outbreak.

## Discussion

The central implication of our analysis is that recent human Andes virus outbreaks are consistent with independent emergences from long-standing, geographically structured reservoir diversity, rather than as a single recently emerged human-transmissible lineage or a repeated, obvious adaptive transition immediately before detection. Across five recorded human outbreak lineages, viruses involved in person-to-person transmission were embedded within broader ANDV diversity sampled from humans and rodents and arose in distinct phylogenetic and regional backgrounds. Our analyses place these observations in a comprehensive segment-linked dataset and a common cross-segment molecular dating framework that retains incompletely sampled isolates. The available genomes suggest that multiple reservoir-associated lineages may already possess some capacity to generate limited human transmission chains. Isolated human infections interspersed among rodent-derived and outbreak-associated diversity further argue against a simple dichotomy between intrinsically non-transmissible reservoir viruses and recently adapted human-transmissible lineages.

Our analysis also addresses a broader methodological challenge in the evolutionary study of segmented viruses. Historical datasets frequently contain incomplete segment sets, inconsistent accession linkage, and different taxon compositions among segment alignments. Restricting analyses to observed segment sequences can discard known relationships among sampled viruses and destabilise inference in sparsely represented segments. Our linkage-informed representation retained real sampled taxa without imputing nucleotide states and allowed isolate identity, sampling time, and segment-supported phylogenetic placement to be represented more consistently across segment-specific time trees. This was particularly valuable for the L segment, where it stabilised inference and recovered temporal model support in a dataset that was unstable using observed sequences alone. It also improved cross-segment comparability for the Chile 2024 lineage: because its closest comparator lacked an observed L segment, the observed-only analysis produced a substantially older time to the most recent common ancestor (tMRCA), whereas linkage-informed representation brought the L-segment estimate into much closer agreement with S and M. The approach is intentionally conservative because it does not account for uncertainty in the constrained topology or reconstruct missing nucleotide states and therefore cannot establish whether an unobserved segment was reassorted. Its benefit should therefore be interpreted as improved use of known linkage and sampling information rather than automatically more accurate reconstruction. Used alongside observed-sequence-only analyses, this framework provides a general way to quantify how patchy segment coverage affects molecular dating inference, particularly by assessing its impact on the compatibility of tMRCA estimates across segments.

Our molecular clock analyses indicate that the evolutionary history of ANDV extends substantially beyond the recent human outbreaks through which many lineages were first recognised. Across nested datasets, the more phylogenetically restricted clades generally produced higher substitution rate estimates, whereas inclusion of progressively older reservoir-associated diversity lowered inferred rates, consistent with time-dependent rate variation and strong purifying selection. Our PoW estimates were older than corresponding tip-dated estimates, with broadly overlapping 95% HPDs for the observed and linkage-informed clades 1-4 datasets, but substantially deeper estimates after inclusion of unclassified diversity. The strongest signal came from the M segment, which contained the greatest number of observed divergent unclassified sequences and placed the sampled root at approximately 17,000 years before present, whereas corresponding S- and L-segment estimates were more uncertain because several unclassified taxa were represented by linkage-informed missing-segment entries. Similar discrepancies between recent tip-dated estimates and deeper evolutionary histories have been reported for Puumala and Tula hantaviruses^22^. Together, these results caution against extrapolating short-term evolutionary rates across deep hantavirus timescales and suggest that the lineages underlying recent ANDV outbreaks may have evolved over much longer periods than their recent detection implies.

This distinction matters for how ANDV emergence is interpreted. The observation of person-to-person spread naturally raises concern that a lineage has acquired new mutations increasing transmissibility in humans. However, our analyses provide little support for a repeated, recent adaptive transition. Branches leading to human outbreaks were dominated by synonymous change, relaxed clock analyses did not identify consistent acceleration on outbreak lineage ancestral branches, branch-specific selection analyses did not detect reproducible evidence of episodic diversifying selection, and structural mapping did not point to any recurrent changes in antigenic, entry, fusion, or polymerase active-site regions. Although several lineage-specific amino acid changes occurred in the glycoprotein or polymerase and remain plausible candidates for experimental follow-up, they affected different sites and structural environments, were generally conservative, and did not converge on a shared functional mechanism. We therefore interpret these substitutions as candidate lineage-specific changes rather than evidence of a common adaptive transition.

An alternative hypothesis is that the capacity for limited human-to-human transmission is an existing property of parts of ANDV diversity, rather than a newly acquired feature of each outbreak lineage. This would imply that outbreaks occur when a reservoir lineage with permissive viral properties enters a favourable ecological and epidemiological context: close contact, delayed diagnosis, household or healthcare transmission, or host factors that allow unusually successful infection^9^. The fact that multiple phylogenetically distinct lineages have caused isolated human infections or limited transmission clusters, yet none has established sustained circulation in humans, is consistent with this interpretation. If a single recent mutation were sufficient to create a stably human-transmissible virus, repeated emergence might be expected to produce more consistent genomic signatures or more persistent transmission. Instead, the key determinant of outbreak risk may lie as much in repeated opportunities for old reservoir lineages to enter settings where limited human transmission becomes possible as in recent adaptive genetic changes in the virus. Limited transmission is therefore likely to reflect an interaction between pre-existing viral properties and contingent ecological, epidemiological, and host conditions.

We also saw limited diversity within the current MV Hondius 2026 outbreak. Across the three genomes analysed here, including paired blood and nasal samples from the same individual, we observed only a single consensus-level difference: the synonymous L-segment mutation L:C2144T. This is also consistent with an independent raw-read-based analysis of additional outbreak sequences, which reported only one further L-segment mutation among samples not included in this study^21^. Our within-host analysis nevertheless identified additional low-frequency variation shared between paired blood and nasal samples, including nonsynonymous variants in the nucleocapsid protein and polymerase. Several variable positions also overlapped sites reconstructed as substitutions along outbreak-lineage ancestral branches. This recurrence across within-host and phylogenetic timescales does not establish adaptation, but it identifies a small set of sites that may be mutationally accessible, selectively tolerated, or functionally relevant. Importantly, none of these variants showed evidence of rising to fixation during the sampled outbreak, supporting the broader conclusion that the MV Hondius transmission chain was not accompanied by an obvious recent adaptive sweep.

Our results also refine what should be tested experimentally. Rather than asking only whether each recent outbreak lineage recently acquired a single transmission-enhancing mutation, future work should ask whether different ANDV clades vary in baseline phenotypes relevant to spillover and person-to-person spread. These include glycoprotein-mediated entry, particle stability, replication efficiency, innate immune antagonism, tissue tropism, shedding, and environmental persistence. The candidate mutations identified here provide a focused starting point, but the broader hypothesis is that phenotypic differences among ANDV lineages may have accumulated deeper in reservoir evolution, before the recent human outbreaks sampled to date.

The geographic patterning of the closest sampled relatives also has direct implications for surveillance. The MV Hondius 2026 lineage is linked to a cross-border southern Chile-northern Patagonia assemblage, the Epuyén and Argentina 2014 lineages to Río Negro-and Chubut-associated diversity, and the Chile 2024 lineage to Neuquén-associated diversity. These patterns are consistent with strong geographic structure within ANDV, but we note that exact phylogeographic reconstruction of the closest ancestor remains challenging given sparse reservoir sampling and incomplete segment sequencing which can dominate uncertainty in timing and immediate ancestry of each outbreak. Surveillance should therefore not focus only on descendants of previously recognised outbreak viruses, because the relevant risk appears to be distributed across incompletely sampled regional reservoir diversity.

In summary, our results suggest that recent human ANDV outbreaks are not the product of obvious, repeated adaptive bursts immediately before detection, but rather sporadic emergences from old, structured, and incompletely sampled reservoir lineages. By integrating new MV Hondius sequence data with a comprehensive segment-linked collection that retains partial and incomplete genomes, the study provides both a broader empirical reconstruction of ANDV diversity and a linkage-informed missing-segment framework for molecular dating of patchy segmented datasets. Systematic temporal-signal testing, time-dependent rate analysis, and PoW modelling place recent outbreaks against a much deeper reservoir-associated history, while substitution reconstruction, selection analysis, branch-rate estimation, structural mapping, and within-host analysis identify no common recent adaptive transition preceding person-to-person transmission. This shifts the question from whether each outbreak lineage newly adapted to humans to which lineage-level phenotypes evolved during deeper ANDV history and which ecological, epidemiological, and host conditions occasionally permit human transmission without sustained circulation.

## Author Contributions

C.S., S.Ö., S.L. and M.G. wrote the original draft. C.S., S.Ö., R.S., A.L., J.A., D.S., L.Z., K.L., A.K., T.B., S.L. and M.G. reviewed and edited the manuscript. C.S., S.Ö., S.L., R.S., A.L., L.Z. and M.G. performed the analyses. J.A. and D.S. generated primary sequence data from the MV Hondius outbreak. C.S., S.Ö., S.L. and M.G. conceived and designed the study. M.G. supervised the work.

## Data Availability

Raw-read pathogen data files are available under BioProject PRJNA1470694 and BioSample IDs SAMN60423882 and SAMN60696784. Curated pathogen alignments, sequence metadata, BEAST XML files, posterior trees, ancestral reconstructions, code and other associated data can be accessed from the following Github repository: https://github.com/spyros-lytras/hantavirus_molevo. Selection analysis and associated dashboards can be accessed from: https://github.com/aglucaci/hantavirus_selection_analysis. Public sequences used in this study are listed with accession numbers, source databases and submitter information in Supplementary Table 1.

https://github.com/aglucaci/hantavirus_selection_analysis

## Acknowledgments

This work was supported by the Wellcome Trust Early-Career Award (grant number 309205/Z/24/Z) to MG. CS was supported by the Oxford-Moh Family Foundation Global Health Scholarship for her DPhil. KL was supported by the Li Ka Shing foundation award and Wellcome Trust grant (grant number 227438/Z/23/Z). TAB is supported by Medical Research Council grants MR/V031635/1 and MR/S007555/1. The Centre for Human Genetics was supported by the Wellcome grant 203141/Z/16/Z. The research was supported by the Wellcome Trust Core Award grant number 203141/Z/16/Z, with funding from the NIHR Oxford BRC. SL is supported by the ANR Choose France for Science grant ANR-25-CFFS-0055.

We would also like to thank all colleagues involved in valuable discussions on the www.virological.org online forum.

## Supplementary Figure legends

**Supplementary Figure 1. *Orthohantavirus andesense* L segment species maximum-likelihood phylogeny.** The Sin Nombre virus L segment is used as the outgroup for rooting the tree. ANDV clade annotations are consistent with those in Figure 1A. The inset at the bottom left of the figure represents the zoomed in ANDV clade of the full tree, human outbreak sequences are highlighted in red and ultrafast bootstrap support values are shown above branches. Scales represent nucleotide substitutions per site.

**Supplementary Figure 2. *Orthohantavirus andesense* S segment species maximum likelihood phylogeny.** The Sin Nombre virus S segment is used as the outgroup for rooting the tree. ANDV clade annotations are consistent with those in Figure 1A. The inset at the bottom left of the figure represents the zoomed in ANDV clade of the full tree, human outbreak sequences are highlighted in red and ultrafast bootstrap support values are shown above branches. Scales represent nucleotide substitutions per site.

**Supplementary Figure 3. *Orthohantavirus andesense* M segment species maximum likelihood phylogeny.** The Sin Nombre virus M segment is used as the outgroup for rooting the tree. ANDV clade annotations are consistent with those in Figure 1A. The inset at the bottom left of the figure represents the zoomed in ANDV clade of the full tree, human outbreak sequences are highlighted in red and ultrafast bootstrap support values are shown above branches. Scales represent nucleotide substitutions per site.

**Supplementary Figure 4. Relaxed clock branch-rate analyses of outbreak-associated clades.** Uncorrelated lognormal relaxed clock analyses across all segments for the five human outbreak-associated lineages, when available. Branches in each time-scaled phylogeny are coloured according to their median evolutionary rate estimates (s/s/y), with blue indicating lower rates and red indicating higher rates. Node labels denote posterior support values, and only values >0.5 are displayed. **(A)** S segment. Evolutionary rate estimates for stems leading to human outbreaks were 2.0x10^-4^ (95% HPD: 8.1x10^-5^–4.6x10^-4^) s/s/y for MV Hondius 2026, 1.4x10^-4^ (95% HPD: 4.8x10^-5^–2.4x10^-4^) s/s/y for Chile 2024, 1.8x10^-4^ (95% HPD: 4.7x10^-5^–7.3x10^-4^) s/s/y for Epuyén 2018/2019 and 1.9x10^-4^ (95% HPD: 4.7x10^-5^–8.6x10^-4^) s/s/y for Argentina 2014. **(B)** M segment. Evolutionary rate estimates for stems leading to human outbreaks were 1.7x10^-4^ (95% HPD: 5.1x10^-5^–3.4x10^-4^) s/s/y for MV Hondius 2026, 2.2x10^-4^ (95% HPD: 1.2x10^-4^–3.4x10^-4^) s/s/y for Chile 2024, 4.0x10^-4^ (95% HPD: 8.4x10^-5^–1.5x10^-3^) s/s/y for Epuyén 2018/2019, 1.4x10^-4^ (95% HPD: 5.1x10^-5^–2.6x10^-4^) s/s/y for Argentina 2014, and 5.1x10^-4^ (95% HPD: 1.1x10^-4^–1.4x10^-3^) s/s/y for Chile 2011. **(C)** L segment. Evolutionary rate estimates for stems leading to human outbreaks were 9.6x10^-4^ (95% HPD: 2.8x10^-4^–2.6x10^-3^) s/s/y for MV Hondius 2026, 1.0x10^-3^ (95% HPD: 2.9x10^-4^–2.3x10^-3^) s/s/y for Chile 2024, 6.7x10^-4^ (95% HPD: 7.6x10^-5^–4.11x10^-3^) s/s/y for Epuyén 2018/2019, and 2.2x10^-4^ (95% HPD: 4.8x10^-5^–4.7x10^-4^) s/s/y for Argentina 2014.

**Supplementary Figure 5. Shared within-host variants in paired MV Hondius blood and nasal samples.** Frequencies of filtered iSNVs in paired blood (XDHK) and nasal (XDJH) samples from the same individual, faceted by genome segment. Thresholds for inclusion are minimum 5% iSNV frequency, and minimum 2 reads supporting iSNV call. Points show variant frequency and stems are colored shaded by log10 read depth. Shapes indicate codon position. Coordinates are numbered relative to the Andes virus reference sequences (NC_*003466.1,* NC_*003467.2,* NC_*003468.2*). Shared variants detected in both sample types include S:909, S:916, M:1164, L:407 and L:3561.

**Supplementary Figure 6. Posterior estimates of tip dates for samples lacking exact collection dates.** Results are shown for the M-segment clades 1-4 analysis under a strict molecular clock and coalescent constant population size model. In each panel, red dashed lines indicate the lower and upper bounds of the uniform prior distribution applied for each tip. Estimates are expressed in years before the most recent sample (2026-05-06). See Supplementary Figures 7 and 8 for the S and L segments, respectively.

**Supplementary Figure 7. Sequences with uncertain collection dates and assigned sampling-date priors in the S segment.** Results are shown for the S-segment clades 1-4 analysis under a strict molecular clock and coalescent constant population size model. In each panel, red dashed lines indicate the lower and upper bounds of the uniform prior distribution applied for each tip. Estimates are expressed in years before the most recent sample (2026-05-06).

**Supplementary Figure 8. Sequences with uncertain collection dates and assigned sampling-date priors in the L segment.** Results are shown for the L-segment clades 1-4 analysis under a strict molecular clock and coalescent constant population size model. In each panel, red dashed lines indicate the lower and upper bounds of the uniform prior distribution applied for each tip. Estimates are expressed in years before the most recent sample (2026-05-06).

## Supplementary Table legends

**Supplementary Table 1. Sequence metadata and inclusion status.** Full metadata for all sequences used in the analysis, including accession, segment, host, country, collection date, source database, submitter or source study, clade assignment, inclusion/exclusion status and analysis subset.

**Supplementary Table 2. Matched Accession IDs across all segments for the same sample/isolate.** Isolate name, country, host, collection date and accessions corresponding to each segment are provided.

**Supplementary Table 3. Sequences with uncertain collection dates and assigned sampling-date priors.** Sequences lacking exact collection dates, inferred sampling intervals from source publications, and uniform priors used in molecular clock analyses.

**Supplementary Table 4. BETS run effective sample size (ESS).** Log-likelihood values and Bayes factor (BF) reported for different sets of data comparing heterochronous (het) and isochronous (iso) models including/excluding starting tree (incl/excl). All datasets were run for 200M iterations with 20 steps, datasets indicated with an asterisk failed at 20 steps and were re-run with 10 steps. Datasets which have BF below 5 have been considered to show lack of evidence of temporal signal (see Methods) and have been indicated. Results are shown for the ‘Observed’ sequences under a strict clock, ‘Observed’ sequences under a relaxed clock, and ‘All’ (linkage-informed) sequences under a strict clock, annotated accordingly at the top row of each sub-table.

**Supplementary Table 5. Inferred clock rate estimates.** Posterior mean, median and 95% highest posterior density intervals for substitution rate estimates across segments, clade compositions, and scenarios. Unless stated otherwise, estimates are inferred under a strict molecular clock model. All estimates are given as substitutions/site/year. Asterisks next to the dataset name indicate analyses for which BETS did not support the presence of a temporal signal.

**Supplementary Table 6. Sequences included in the two-per-outbreak sensitivity analyses.** Accession IDs for the reduced “2pO” datasets, in which the two earliest available samples from each outbreak cluster were retained to assess the effect of dense sampling of near-identical outbreak genomes on molecular clock estimates.

**Supplementary Table 7. Stem-branch mutation reconstruction.** Nucleotide and amino acid mutations within the coding regions inferred on branches leading to human outbreak-associated lineages, with synonymous/nonsynonymous classification, protein annotation. Genome nucleotide positions are with respect to reference for each segment-S: NC_003466.1,M: NC_003467.2 and L: NC_003468.2.

**Supplementary Table 8. Within-host variants in MV Hondius blood and nasal samples.** Within-host single-nucleotide variants identified in the paired MV Hondius blood and nasal samples. Variants were included if they had a minimum iSNV frequency of 5% and were supported by at least two reads. Yellow highlighting indicates iSNV positions that were also reconstructed as mutations on one or more of the five outbreak stem branches. Reference genomes are S: NC_003466.1, M: NC_003467.2 and L: NC_003468.2. Sample identifiers are XDHK: PP_006XDHK.5 and XDJH: PP_006XDJH.5.

## Methods

### Sequence generation and consensus calling

Two primary genome sequences from the current MV Hondius-associated Andes virus outbreak were generated from paired clinical samples collected from the same individual on the same day: a whole-blood sample (XDHK) and a nasopharyngeal swab sample (XDJH). Laboratory processing followed the tiled-amplification approach described previously for Andes virus sequencing^23^, using Oxford Nanopore MinION sequencing.

Viral RNA was extracted using the QIAamp Viral RNA kit (Qiagen). Reverse transcription was performed using SuperScript IV reverse transcriptase (ThermoFisher Scientific), following the manufacturer’s recommendations, with 8 μl of extracted RNA as input. PCR amplification was carried out using Phusion U Multiplex Master Mix (ThermoFisher Scientific), with 2 μl of cDNA template in 25 μl reactions. Cycling conditions were 98°C for 2 min, followed by 45 cycles of 98°C for 30 s and 65°C for 5 min, with a final extension at 72°C for 10 min.

PCR products were purified using 0.8× MagMAX PureBind magnetic beads (ThermoFisher Scientific), washed twice with 80% ethanol, and eluted in 20 μl of 1 mM Tris pH 8.0. Purified amplicon pools were quantified using the Qubit Broad Range dsDNA 1× assay kit (ThermoFisher Scientific) and combined in equal proportions where possible. Pooled amplicons were prepared for sequencing using the Native Barcoding Kit V14 (SQK-NBD114.24; Oxford Nanopore Technologies) and sequenced on a MinION Mk1D device using a FLO-MIN114 flow cell.

Raw sequence data were analysed using nf-ViralMutations (https://github.com/phac-nml/nf-ViralMutations; commit 8675768). Reads were aligned to Andes virus reference sequences and processed to generate segment-specific consensus sequences and base-frequency summaries. Human and other off-target reads were removed before downstream analysis. Consensus positions were masked when read support was below 20. In the event that two or more mutations combine to more than 70% (e.g. one mutation at 35% and one at 40%), then the IUPAC ambiguous base of the mutations is called.

### Sequence and corresponding data curation

We downloaded all nucleotide sequences classified in the species *Orthohantavirus andesense* (taxid: 1980456) available on the NCBI Virus database as of the 26^th^ of May 2026. Additionally, we included the first genome of the recent MV Hondius 2026 outbreak sampled in Switzerland (Pathoplexus: PP_006WBLH.2, GenBank: L - PZ385161.1, M - PZ385162.1, S - PZ385163.1, ANDV/Switzerland/Hu-3337/2026) as well as two sequences from the same patient of the outbreak generated as part of this study (Pathoplexus: PP_006XDJH.5 and PP_006XDHK.5). Lastly, the reference genome of Sin Nombre virus (GenBank: L - NC_077671.1, M - NC_077670.1, S - NC_077669.1) was included in the analysis as an outgroup.

Metadata were harmonised across sources, including accession, segment, host, country, region or province where available, collection date, source study, and data-use status. Sequences were excluded from downstream analysis if they were derived from experimental passage (**Supplementary Table 1**). For records without exact collection dates, we consulted the source publications and assigned bounded sampling intervals. These intervals were incorporated as uniform tip-date priors in BEAST analyses (**Supplementary Table 3, Supplementary Figure 6-8**).

### Multiple sequence alignment

Before constructing the sequence alignments, partial M segment sequences covering different parts of the same virus genome segment (Gc/Gn) were manually concatenated based on their isolate names and information available in their original publication. Nucleotide sequences of each segment were then aligned using mafft v7.505 --localpair option^24^. These alignments were manually trimmed to their coding sequences and translated using BioEdit^25^ and realigned on the amino acid level with mafft --localpair. The resulting amino acid alignments were converted to their corresponding codon alignments using pal2nal v14^26^, which were used for all phylogenetic and selection analysis described below. Coding-region coordinates were defined relative to the Andes virus reference genome^2^ as follows: S segment, nucleotides 43-1329; M segment, nucleotides 52-3468; and L segment, nucleotides 36-6497.

### Maximum-likelihood phylogenetics and clade definition

Maximum-likelihood trees were inferred separately for the S, M and L segment codon alignments using IQ-TREE v2.3.5^27^. ModelFinderPlus was used to identify best-fitting nucleotide substitution models^28^, and branch support was assessed with 10,000 ultrafast bootstrap replicates^29^. The species tree was separated into ANDV-like background and ANDV, focusing on the four ANDV clades defined consistent with Bellomo et al^6^. Sequences within ANDV diversity that were not monophyletic with any of the four clades were annotated as “unclassified” **(Supplementary Figures 1-3)**. Trees were visualised and annotated in FigTree (https://github.com/rambaut/figtree).

For each outbreak-associated lineage, we compared the reconstructed sequence at the closest sampled common ancestor with the reconstructed sequence on the branch leading to the outbreak cluster. Nucleotide changes were annotated by segment, genomic coordinate, codon, codon position, and amino acid change using custom R script.

### Reconstruction of linked genome-segment datasets

ANDV sequence records were linked across the S, M, and L genome segments using isolate names, accession metadata, source publications, sampling location, host, and collection date. Segment linkages were accepted only where the records could be assigned to the same sampled virus with high confidence. The resulting linkage table is provided in **Supplementary Table 2.**

For a sampled virus with an observed sequence for at least one genome segment but no available sequence for another segment(s), we created a linkage-informed missing-segment entry. This entry consisted entirely of N characters and matched the length of the corresponding segment alignment. It was assigned the same isolate identifier, host metadata, and sampling date as the linked observed segment. No nucleotide state was imputed.

The placement of each missing-segment taxon was constrained to the clade supported by its observed linked segment, while relationships within that clade remained estimated from the segment-specific data. This approach assumes that the missing segment has not undergone reassortment that would place it outside the clade supported by the observed linked segment. Although this assumption cannot be tested directly for an unsequenced segment, reassortment was limited across the sampled ANDV phylogenies and was not detected among the outbreak lineages or their closest comparator sequences (i.e., non-outbreak sequences that provide phylogenetic context for placing and dating outbreak lineages within broader ANDV diversity). We therefore considered clade-level placement to be a reasonable working assumption, while retaining observed-sequence-only analyses as sensitivity analyses.

Because all-N sequences are uninformative with respect to nucleotide character likelihood, these entries contributed no direct sequence evidence. Their purpose was to preserve known taxon linkage, sampling time, and topological representation across segment-specific analyses.

### Molecular clock analyses

Time-scaled phylogenies were inferred using BEAST 2 v2.6.3^30^. Analyses were performed separately for each genome segment and for pre-defined outbreak-associated clade subsets containing human outbreak lineages and their nearest available human- or rodent-derived comparator sequences.

For the primary dated analyses, we focused on ANDV clades 1-4 and five outbreaks with documented person-to-person transmission where sequence data were available: MV Hondius 2026, Chile 2011, Argentina 2014, Epuyén 2018/19, and Chile 2024. M-segment data were available for all five, whereas S and L analyses included four because comparable sequence data were unavailable for Chile 2024. EU241684.1 (S) and EU241643.1-EU241657.1 (M) had discordant or weakly supported placements between maximum-likelihood and BEAST trees. In the BEAST time trees, these sequences fell outside clades 1-4 and were treated as unclassified in downstream analyses.

Primary analyses used a strict molecular clock and constant size coalescent prior. Complementary uncorrelated lognormal relaxed clock analyses were performed for S, M, and L to compare branch-specific rates across the full clades 1-4 dataset, the MV Hondius-related clades 1+3 subset, and the Epuyén-related clade 2 subset (**Supplementary Tables 4 and 5**). Strict clock analyses converged consistently and were used for the main node-date estimates. Relaxed clock analyses were used for branch-rate comparisons and sensitivity analyses rather than for the primary dates.

Analyses used an HKY substitution model. Uniform priors ranging from 10^-12^ to 10^-2^ substitutions/site/year were assigned to strict clock rate and relaxed clock mean parameters. MCMC chains were run for 10-400 million iterations, with the first 10% discarded as burn-in. Convergence and mixing were assessed using Tracer v1.7.2^31^, requiring effective sample size values above 200 for all the parameters included in the model. Maximum clade credibility (MCC) trees with median heights and assuming burn-in of 10% were summarised using TreeAnnotator and visualised using FigTree v1.4.4.

### Closest sampled common ancestors and outbreak-cluster sensitivity analyses

For each outbreak-associated lineage, we identified the posterior node corresponding to the closest sampled common ancestor between the outbreak lineage and its nearest available comparator sequence or comparator clade. Posterior distributions of these node dates were extracted from BEAST posterior trees. These dates were interpreted as closest sampled common ancestor dates, not as direct estimates of spillover timing or the first human infection.

Because several outbreak datasets contained multiple near-identical sequences from the same human transmission cluster, we performed sensitivity analyses to test whether dense outbreak sampling affected coalescent-based node-date estimates. For each outbreak cluster, we repeated primary analyses using a reduced dataset retaining two representative sequences per outbreak where available. Representative sequences were chosen to preserve temporal and genetic diversity within each outbreak while reducing overrepresentation of nearly identical genomes. These analyses were compared with the all-sequence analyses to assess whether inferred closest sampled common ancestor dates were robust to dense sampling of recent outbreak sequences. In parallel, we compared these datasets with the inclusion of linkage-informed missing-segment taxa to assess the effect of incomplete comparator-segment representation on divergence time and rate estimates.

### Bayesian evaluation of temporal signal

Temporal signal was evaluated using Bayesian Evaluation of Temporal Signal (BETS) ^12^. For each segment, heterochronous models incorporating observed sampling dates were compared with otherwise equivalent isochronous models in which all tips shared the same date. Analyses were conducted separately for clade 2, clades 1+3, clades 1-3, and clades 1-4. To assess sensitivity to initial topology, analyses were repeated with and without a starting tree where indicated. We separately ran the same set of tests for datasets containing linkage-informed missing-segment taxa only where observed-only datasets lacked support for temporal signal, for eg. L clade 1-3. For the S segment there were 12 missing samples, 6 for M, and 29 for L segment.

The heterochronous and isochronous models used identical substitution, molecular clock, and tree-prior specifications apart from their treatment of sampling times. Marginal likelihoods were estimated using stepping-stone sampling with 20 path steps, a chain length of 200 million per step, and 10% burn-in unless stated otherwise. Log Bayes factors were calculated as the difference between the log marginal likelihood of the heterochronous and isochronous models. Positive values favoured temporal structure, with values above +5 units interpreted as evidence for presence of temporal signal (see **Supplementary Table 4**).

### Codon-based selection analyses

Selection analyses were performed across local outbreak-associated clades and broader ANDV phylogeny. We estimated site-specific and branch-specific dN/dS and used codon-substitution methods^32^ to test for pervasive negative selection (FEL)^33^, episodic diversifying selection (MEME and FEL)^34^ and branch-specific shifts in selective pressure (aBSREL^35^, RELAX^36^). Outbreak stem branches were labelled a priori to test whether they showed evidence of elevated nonsynonymous evolution relative to background branches.

### Structural mapping of nonsynonymous mutations

Nonsynonymous mutations were mapped to available ANDV or homologous hantavirus protein structures. For the M-segment glycoprotein precursor, residues were interpreted relative to the deposited ANDV glycoprotein structure PDB 9P3I and published domain annotations for the Gn cytoplasmic tail, transmembrane region and putative RNP/N-protein-binding region. For the L-segment RdRp mutation, no ANDV RdRp structure was available, so the mutation was mapped by sequence alignment to Hantaan virus RdRp structures, including the full-length RdRp structure PDB 8P1L and the endonuclease-domain structure PDB 5IZE. Structural context was assessed by residue location, solvent exposure, proximity to annotated functional regions, active sites, transmembrane regions, and unresolved loops and linkers.

### Reconstruction of deeper evolutionary history of ANDV

To reconstruct deeper evolutionary timescales, we applied the Prisoner of War (PoW) model to broader ANDV diversity^8^. The PoW model maps ultrametric genetic-distance trees to time-scaled divergence trees by accounting for the decline in apparent substitution rate over longer evolutionary timescales. This approach is useful when short-term rates estimated from measurably evolving datasets cannot be extrapolated linearly over deeper divergence times because of time-dependent rate variation.

We used posterior substitution rate distributions from the clade 2 dataset shown in **Figure 2A** as the short-term calibration for PoW reconstruction. For each segment, we then generated ultrametric genetic-distance trees for three datasets: ANDV clades 1-4, ANDV clades 1-4 including unclassified sequences, and ANDV clades 1-4 datasets containing linkage-informed missing-segment taxa and unclassified sequences. We then transformed genetic node heights into calendar times using the PoW distance-to-time transformation and obtained branch durations as differences between transformed parent and child node ages.

To propagate uncertainty, we sampled from the posterior distribution of short-term rates and from the posterior distribution of ultrametric distance trees. The resulting time-scaled trees were summarised as maximum clade credibility trees with median heights and assuming burn-in of 10% using TreeAnnotator. We used these reconstructions to estimate divergence times among major ANDV clades, to assess the deeper ancestry of the sampled ANDV lineage, and to compare the timescale of major reservoir-associated clades with the recent human outbreaks through which many lineages were detected.

## Data and code availability

Raw-read data files are available under BioProject PRJNA1470694 and BioSample IDs SAMN60423882 and SAMN60696784. Curated alignments, metadata, BEAST XML files, posterior trees, ancestral reconstructions, code and other associated data can be accessed from the following Github repository: https://github.com/spyros-lytras/hantavirus_molevo. Selection analysis and associated dashboards can be accessed from: https://github.com/aglucaci/hantavirus_selection_analysis.

Public sequences used in this study are listed with accession numbers, source databases and submitter information in **Supplementary Table 1**.

