## supplementary figures for "Molecular evolution of Andes virus lineages leading to recent human outbreaks"

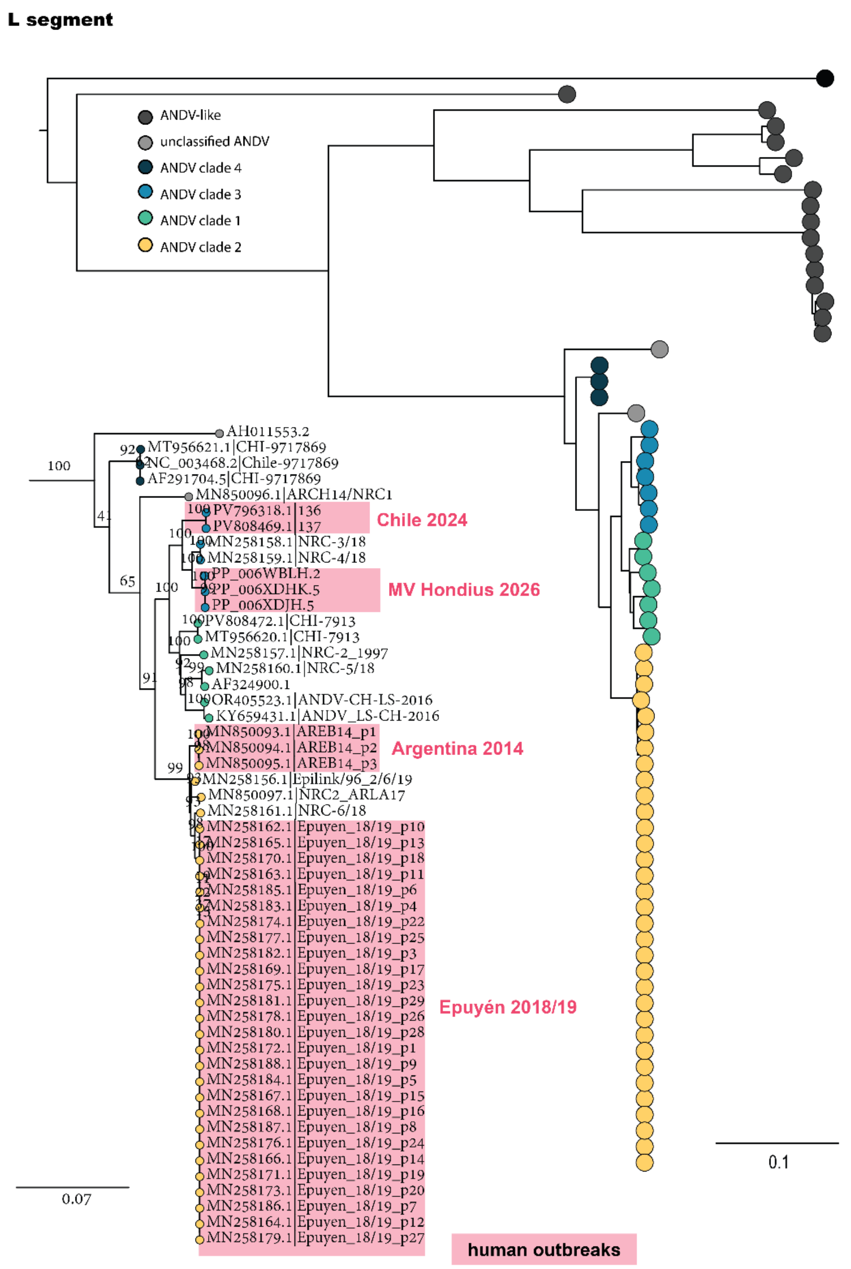


**Supplementary Figure 1. *Orthohantavirus andesense* L segment species maximum-likelihood phylogeny.** The Sin Nombre virus L segment is used as the outgroup for rooting the tree. ANDV clade annotations are consistent with those in Figure 1A. The inset at the bottom left of the figure represents the zoomed in ANDV clade of the full tree, human outbreak sequences are highlighted in red and ultrafast bootstrap support values are shown above branches. Scales represent nucleotide substitutions per site.

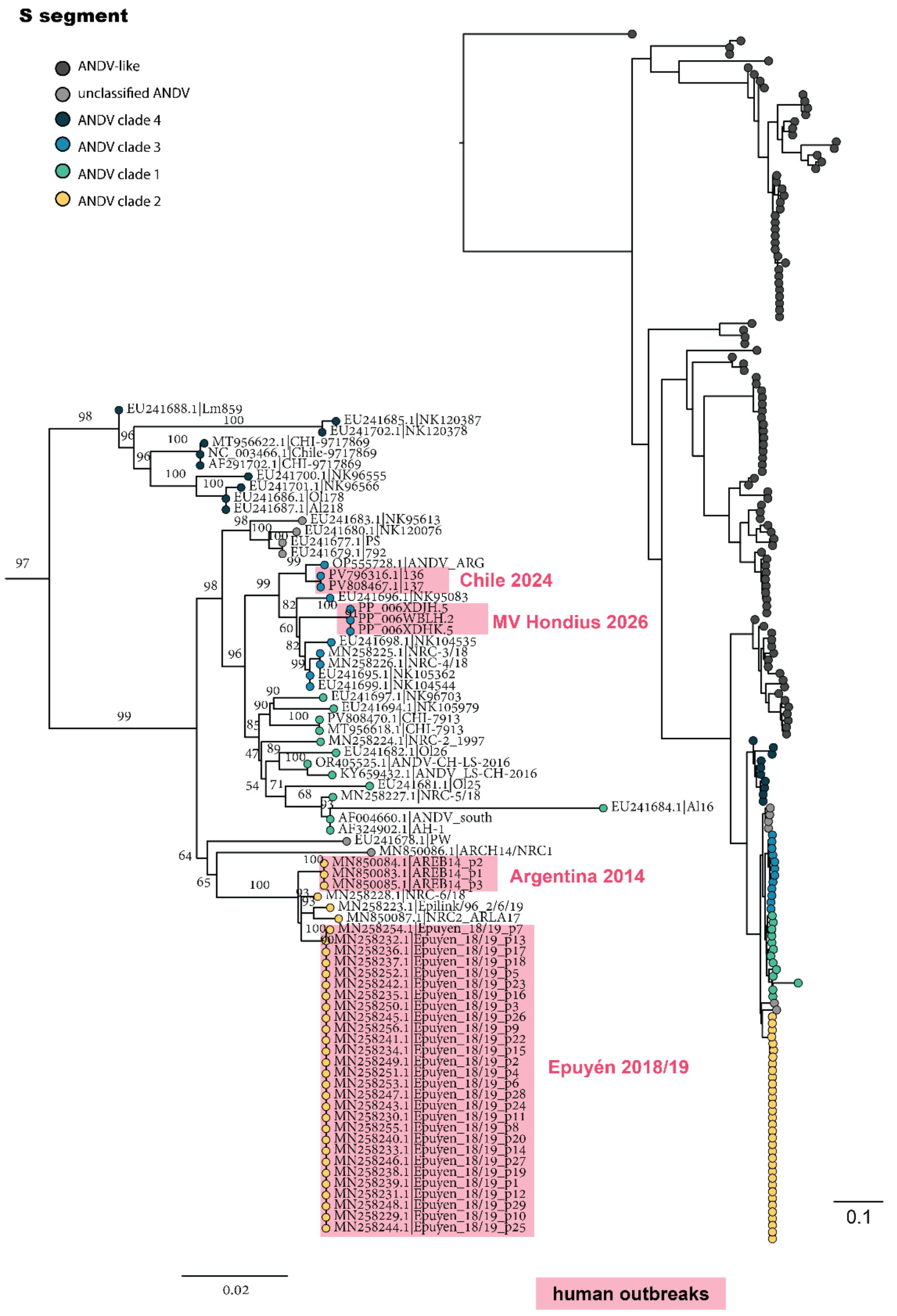


**Supplementary Figure 2.** ***Orthohantavirus andesense*** **S segment species maximum likelihood phylogeny.** The Sin Nombre virus S segment is used as the outgroup for rooting the tree. ANDV clade annotations are consistent with those in Figure 1A. The inset at the bottom left of the figure represents the zoomed in ANDV clade of the full tree, human outbreak sequences are highlighted in red and ultrafast bootstrap support values are shown above branches. Scales represent nucleotide substitutions per site.

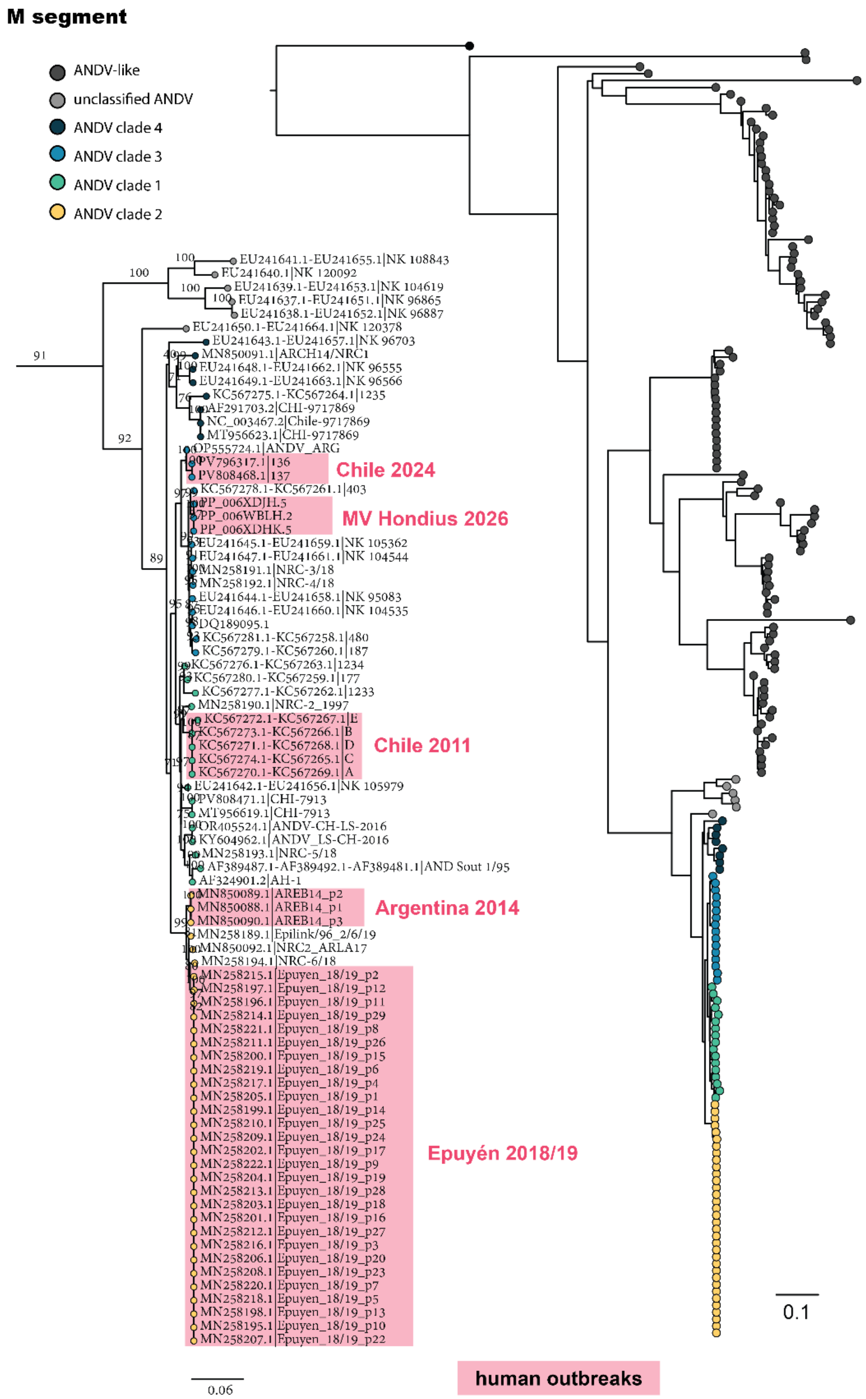


**Supplementary Figure 3.** ***Orthohantavirus andesense*** **M segment species maximum likelihood phylogeny.** The Sin Nombre virus M segment is used as the outgroup for rooting the tree. ANDV clade annotations are consistent with those in Figure 1A. The inset at the bottom left of the figure represents the zoomed in ANDV clade of the full tree, human outbreak sequences are highlighted in red and ultrafast bootstrap support values are shown above branches. Scales represent nucleotide substitutions per site.

### A. S segment

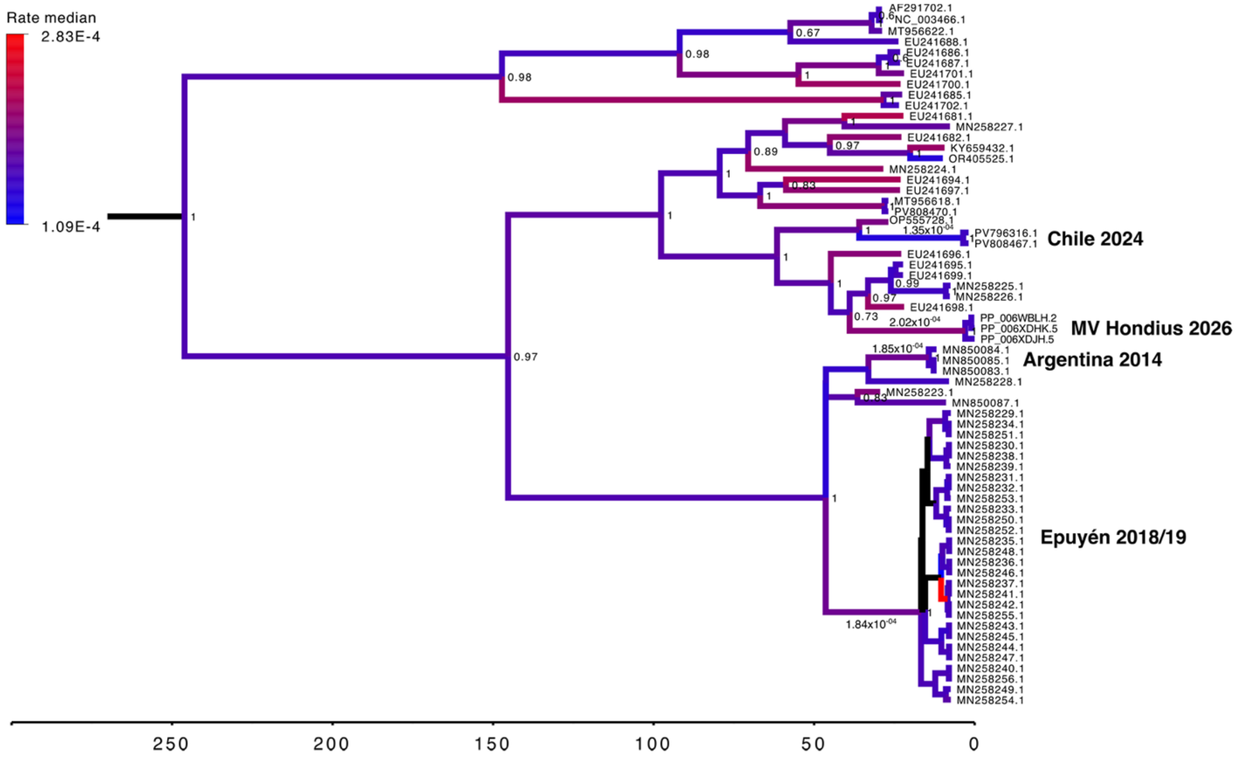


### B. M segment

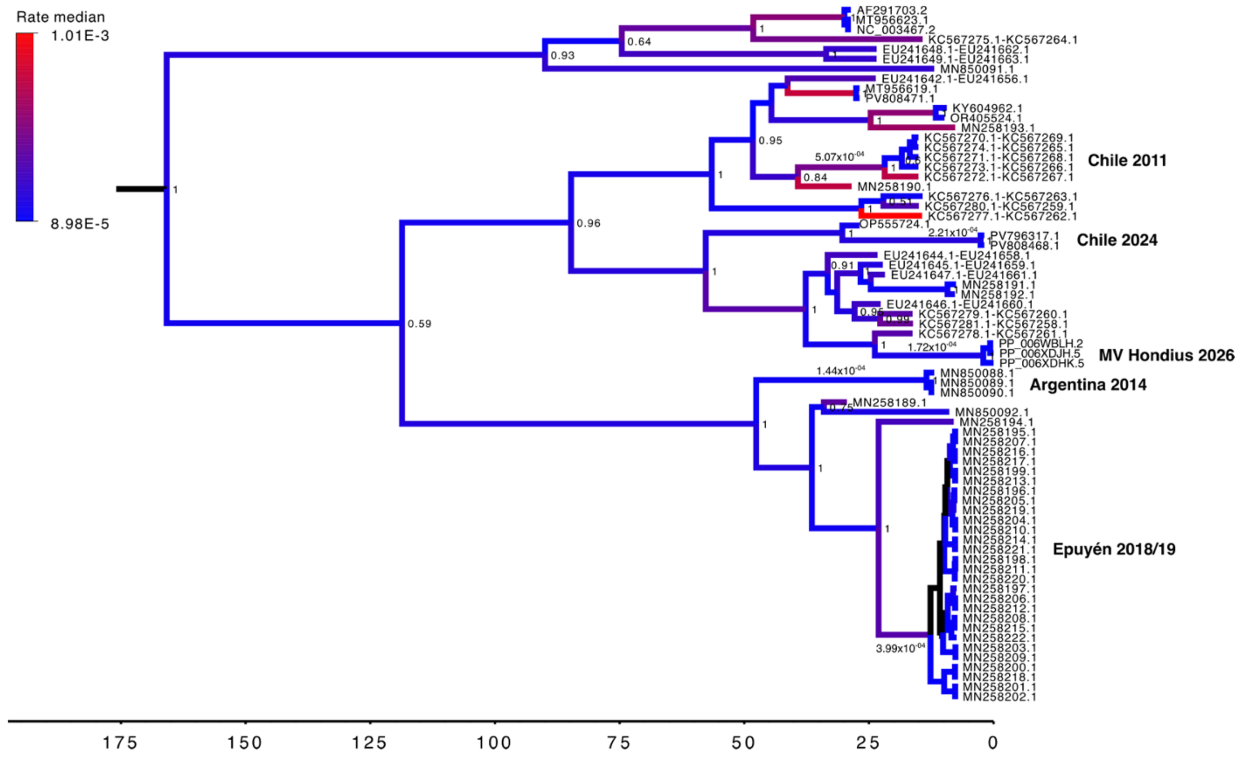

### C. L segment

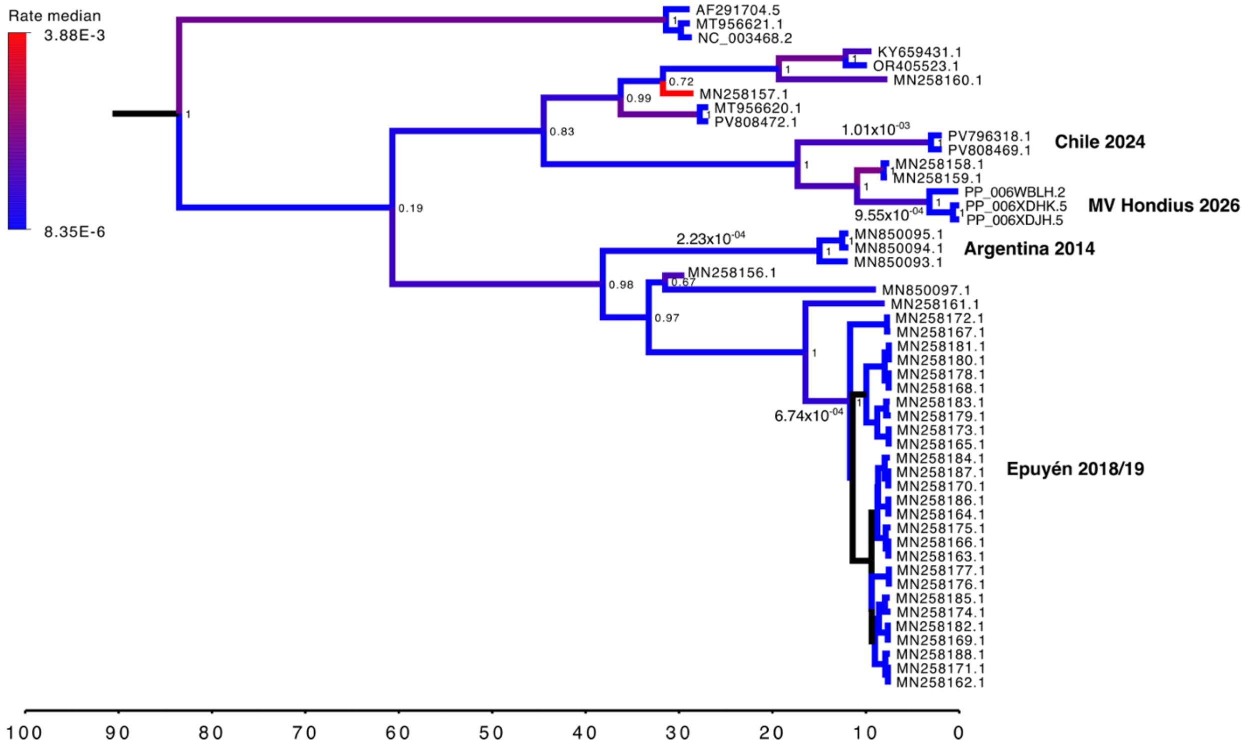


**Supplementary Figure 4. Relaxed clock branch-rate analyses of outbreak-associated clades.**

**Supplementary Figure 4 (cont).** Uncorrelated lognormal relaxed clock analyses across all segments for the five human outbreak-associated lineages, when available. Branches in each time-scaled phylogeny are coloured according to their median evolutionary rate estimates (s/s/y), with blue indicating lower rates and red indicating higher rates. Node labels denote posterior support values, and only values >0.5 are displayed. **(A)** S segment. Evolutionary rate estimates for stems leading to human outbreaks were $2.0x10^{-4}$ (95% HPD: $8.1x10^{-5}$–$4.6x10^{-4}$) s/s/y for MV Hondius 2026, $1.4x10^{-4}$ (95% HPD: $4.8x10^{-5}$–$2.4x10^{-4}$) s/s/y for Chile 2024, $1.8x10^{-4}$ (95% HPD: $4.7x10^{-5}$–$7.3x10^{-4}$) s/s/y for Epuyén 2018/2019 and $1.9x10^{-4}$ (95% HPD: $4.7x10^{-5}$–$8.6x10^{-4}$) s/s/y for Argentina 2014. **(B)** M segment. Evolutionary rate estimates for stems leading to human outbreaks were $1.7x10^{-4}$ (95% HPD: $5.1x10^{-5}$–$3.4x10^{-4}$) s/s/y for MV Hondius 2026, $2.2x10^{-4}$ (95% HPD: $1.2x10^{-4}$–$3.4x10^{-4}$) s/s/y for Chile 2024, $4.0x10^{-4}$ (95% HPD: $8.4x10^{-5}$–$1.5x10^{-3}$) s/s/y for Epuyén 2018/2019, $1.4x10^{-4}$ (95% HPD: $5.1x10^{-5}$–$2.6x10^{-4}$) s/s/y for Argentina 2014, and $5.1x10^{-4}$ (95% HPD: $1.1x10^{-4}$–$1.4x10^{-3}$) s/s/y for Chile 2011. **(C)** L segment. Evolutionary rate estimates for stems leading to human outbreaks were $9.6x10^{-4}$ (95% HPD: $2.8x10^{-4}$–$2.6x10^{-3}$) s/s/y for MV Hondius 2026, $1.0x10^{-3}$ (95% HPD: $2.9x10^{-4}$–$2.3x10^{-3}$) s/s/y for Chile 2024, $6.7x10^{-4}$ (95% HPD: $7.6x10^{-5}$–$4.11x10^{-3}$) s/s/y for Epuyén 2018/2019, and $2.2x10^{-4}$ (95% HPD: $4.8x10^{-5}$–$4.7x10^{-4}$) s/s/y for Argentina 2014.

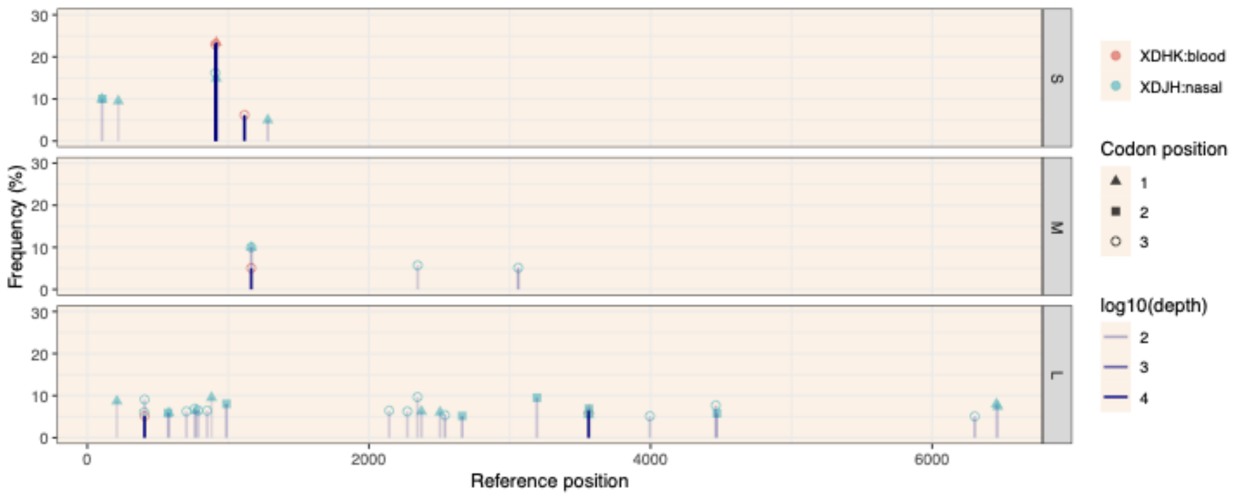


**Supplementary Figure 5. Shared within-host variants in paired MV Hondius blood and nasal samples.** Frequencies of filtered iSNVs in paired blood (XDHK) and nasal (XDJH) samples from the same individual, faceted by genome segment. Thresholds for inclusion are minimum 5% iSNV frequency, and minimum 2 reads supporting iSNV call. Points show variant frequency and stems are colored shaded by log10 read depth. Shapes indicate codon position. Coordinates are numbered relative to the Andes virus reference sequences (NC_*003466.1,* NC_*003467.2,* NC_*003468.2*). Shared variants detected in both sample types include S:909, S:916, M:1164, L:407 and L:3561.

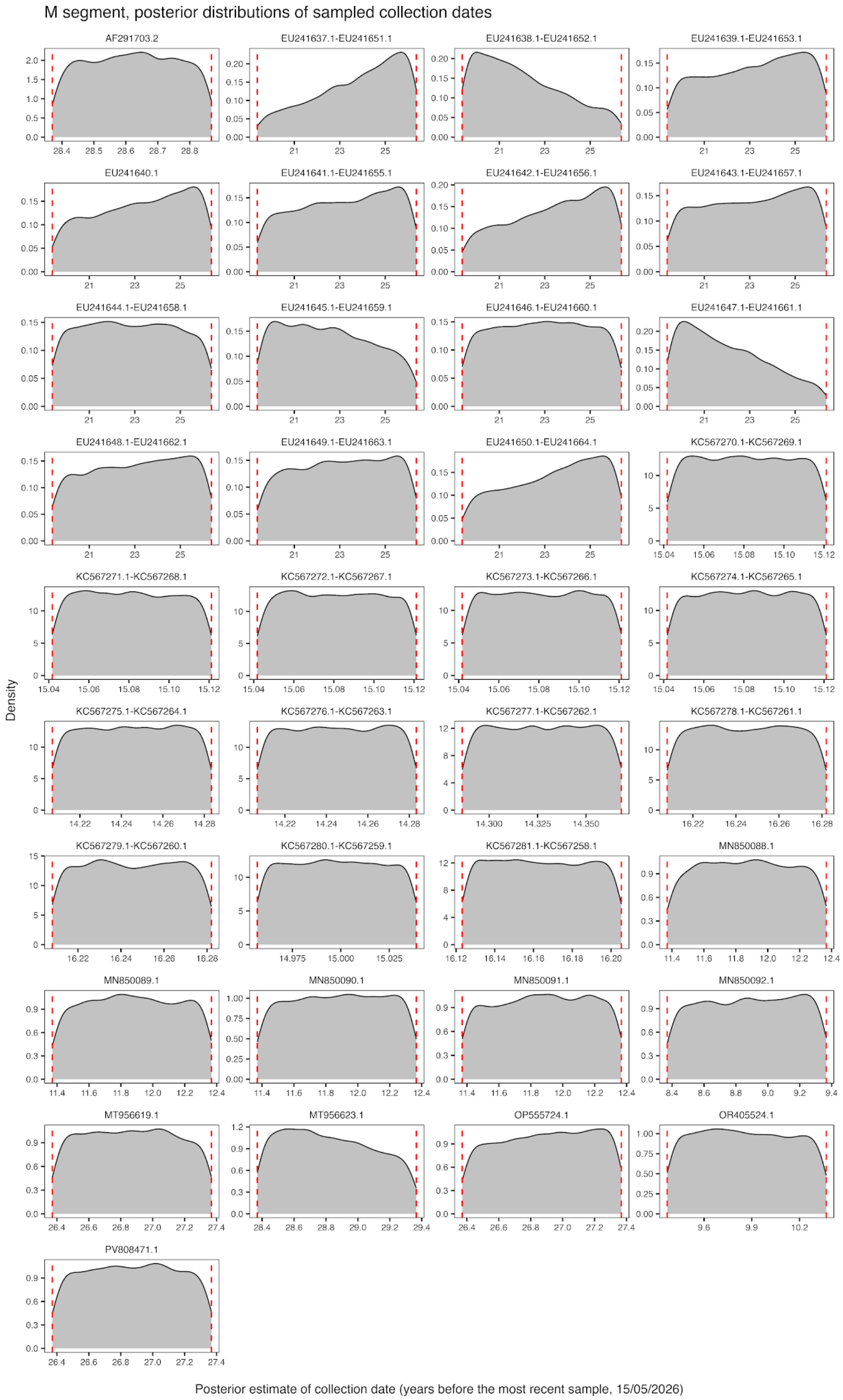


**Supplementary Figure 6. Posterior estimates of tip dates for samples lacking exact collection dates.** Results are shown for the M-segment clades 1-4 analysis under a strict molecular clock and coalescent constant population size model. In each panel, red dashed lines indicate the lower and upper bounds of the uniform prior distribution applied for each tip. Estimates are expressed in years before the most recent sample (2026-05-06). See Supplementary Figures 7 and 8 for the S and L segments, respectively.

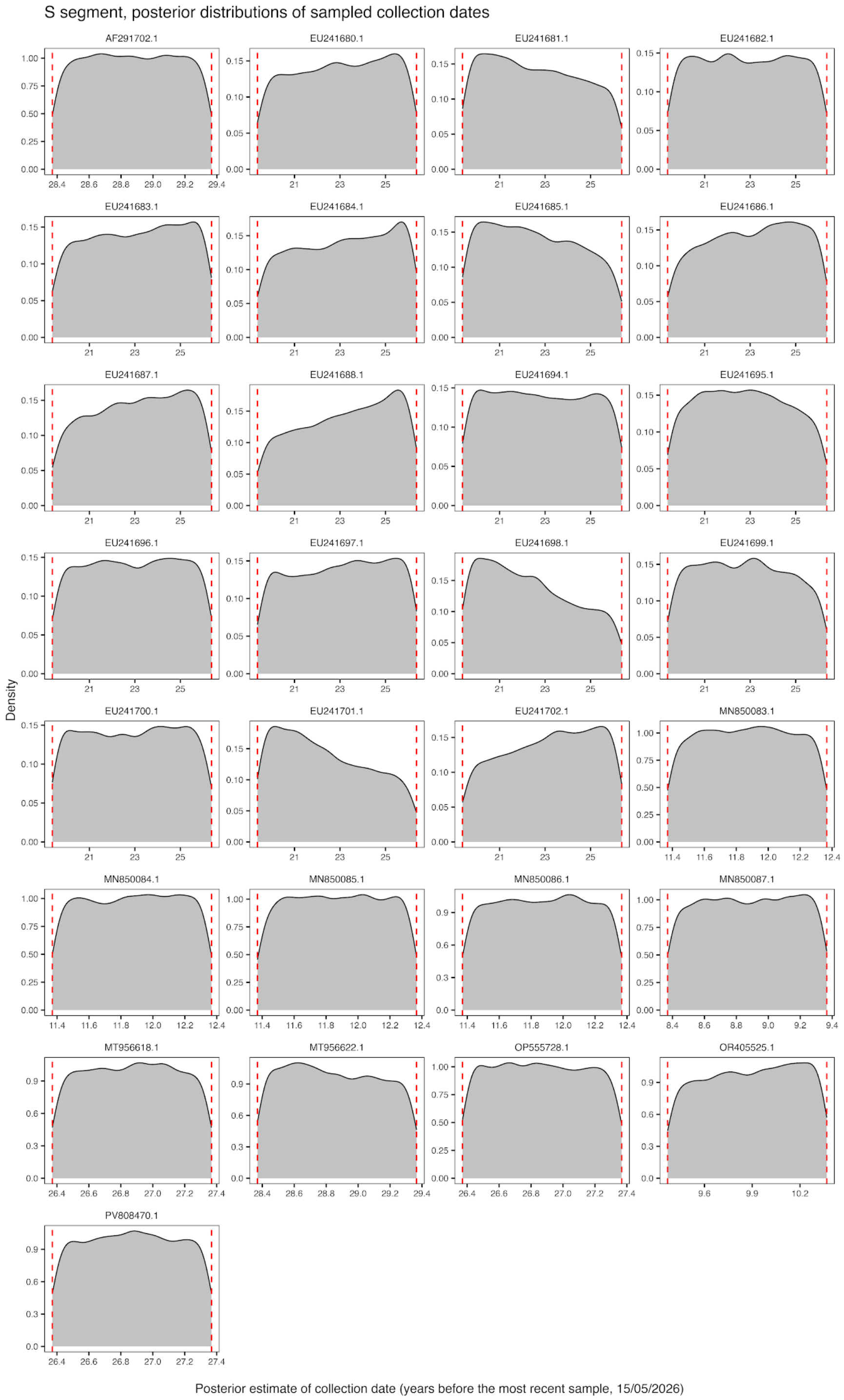


**Supplementary Figure 7. Sequences with uncertain collection dates and assigned sampling-date priors in the S segment.** Results are shown for the S-segment clades 1-4 analysis under a strict molecular clock and coalescent constant population size model. In each panel, red dashed lines indicate the lower and upper bounds of the uniform prior distribution applied for each tip. Estimates are expressed in years before the most recent sample (2026-05-06).

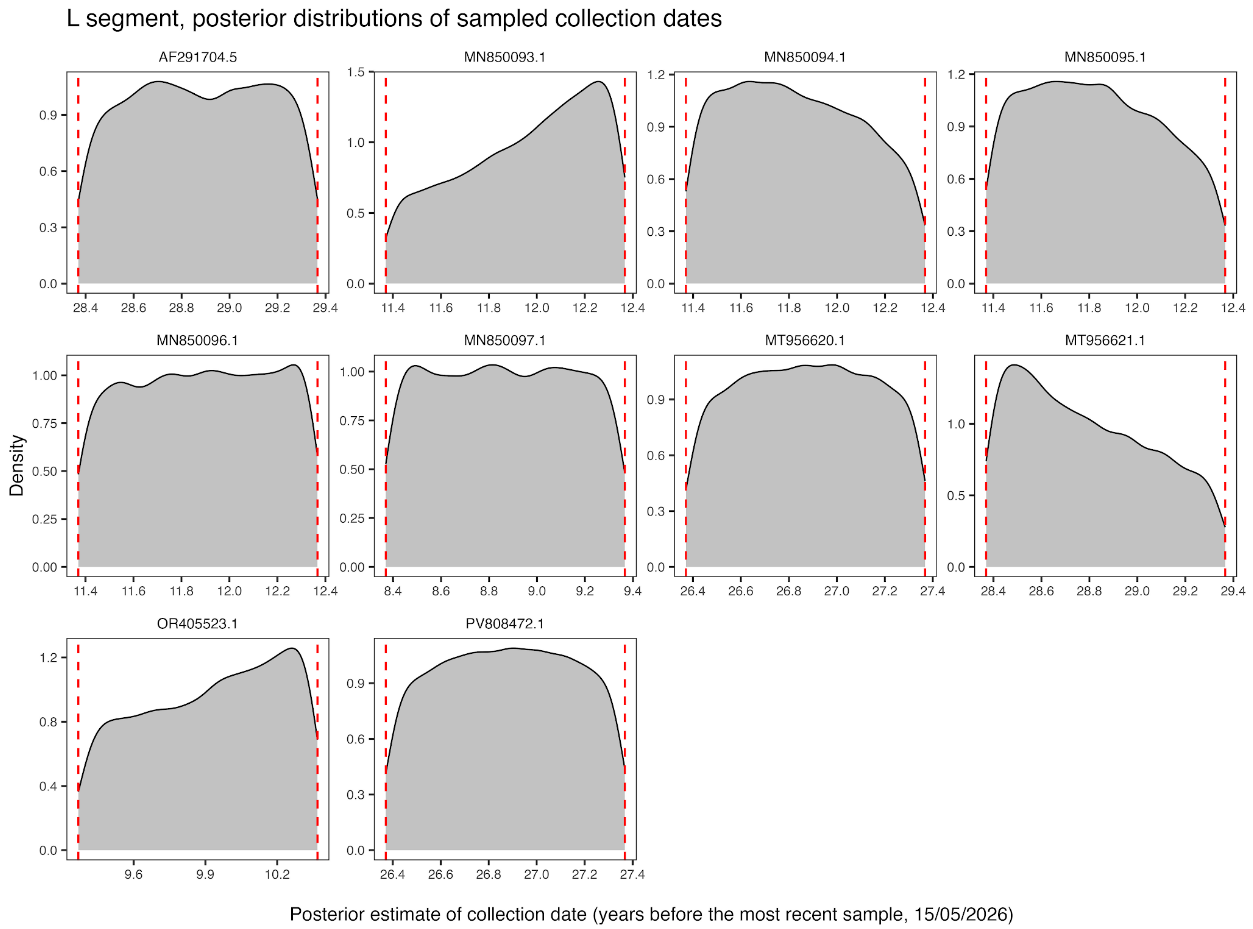


**Supplementary Figure 8**. **Sequences with uncertain collection dates and assigned sampling-date priors in the L segment.** Results are shown for the L-segment clades 1-4 analysis under a strict molecular clock and coalescent constant population size model. In each panel, red dashed lines indicate the lower and upper bounds of the uniform prior distribution applied for each tip. Estimates are expressed in years before the most recent sample (2026-05-06).
